# Immune Cell Densities Predict Response to Immune Checkpoint-Blockade in Head and Neck Cancer

**DOI:** 10.1101/2024.09.10.24313432

**Authors:** Daniel A. Ruiz-Torres, Michael E. Bryan, Shun Hirayama, Ross D. Merkin, Evelyn Luciani, Thomas Roberts, Manisha Patel, Jong C. Park, Lori J. Wirth, Peter M. Sadow, Moshe Sade-Feldman, Shannon L. Stott, Daniel L. Faden

**Affiliations:** Department of Otolaryngology-Head and Neck Surgery, Harvard Medical School, Boston, MA 02115, USA; Massachusetts Eye and Ear, Boston, MA 02118, USA; Massachusetts General Hospital Cancer Center, Boston, MA 02118, USA; Harvard Medical School, 25 Shattuck St, Boston, MA 02115; Department of Pathology, Massachusetts General Hospital, Harvard Medical School, Boston, Massachusetts, USA; Center for Engineering in Medicine and BioMEMS Resource Center, Surgical Services, Massachusetts General Hospital, Harvard Medical School, 114 16th Street, Charlestown, MA 02129, USA; Broad Institute of MIT and Harvard, Cambridge, MA 02142, USA

## Abstract

Immune checkpoint blockade (ICB) is the standard of care for recurrent/metastatic head and neck squamous cell carcinoma (HNSCC), yet efficacy remains low. The current approach for predicting the likelihood of response to ICB is a single proportional biomarker (PD-L1) expressed in immune and tumor cells (Combined Positive Score, CPS) without differentiation by cell type, potentially explaining its limited predictive value. Tertiary Lymphoid Structures (TLS) have shown a stronger association with ICB response than PD-L1. However, their exact composition, size, and spatial biology in HNSCC remain understudied. A detailed understanding of TLS is required for future use as a clinically applicable predictive biomarker.

**Methods:** Pre-ICB tumor tissue sections were obtained from 9 responders (complete response, partial response, or stable disease) and 11 non-responders (progressive disease) classified via RECISTv1.1. A custom multi-immunofluorescence (mIF) staining assay was designed, optimized, and applied to characterize tumor cells (pan-cytokeratin), T cells (CD4, CD8), B cells (CD19, CD20), myeloid cells (CD16, CD56, CD163), dendritic cells (LAMP3), fibroblasts (α Smooth Muscle Actin), proliferative status (Ki67) and immunoregulatory molecules (PD1). Spatial metrics were compared among groups. Serial tissue sections were scored for TLS in both H&E and mIF slides. A machine learning model was employed to measure the effect of these metrics on achieving a response to ICB (SD, PR, or CR).

**Results:** A higher density of B lymphocytes (CD20+) was found in responders compared to non-responders to ICB (p=0.022). A positive correlation was observed between mIF and pathologist identification of TLS (*R^2^= 0.66, p-value= <0.0001*). TLS trended toward being more prevalent in responders to ICB (p=0.0906). The presence of TLS within 100 µm of the tumor was associated with improved overall (p=0.04) and progression-free survival (p=0.03). A multivariate machine learning model identified TLS density as a leading predictor of response to ICB with 80% accuracy.

**Conclusion:** Immune cell densities and TLS spatial location within the tumor microenvironment play a critical role in the immune response to HNSCC and may potentially outperform CPS as a predictor of ICB response.

## Introduction

Head and neck squamous cell carcinoma (HNSCC) is the seventh most common malignancy worldwide and accounts for 4.5% of cancer diagnoses and deaths (1,2). In the United States, 15% of cases present with distant metastasis, for which the 5-year survival rate is 39.3% (3). Immune checkpoint blockade (ICB), with or without chemotherapy, is the standard of care for recurrent or metastatic HNSCC, but only 20% of patients achieve a clinical benefit (4–6). The Combined Positive Score (CPS) is the proportion of all PD-L1 positive cells-including tumor, immune, and stromal cells- among all cells in a single section of tumor tissue. The CPS is calculated manually by a pathologist and is often used to assist with selecting a treatment modality and estimating prognosis (7). However, CPS has limited performance in predicting response to treatment (8). Therefore, an unmet need exists for accurate predictive biomarkers of response to ICB in HNSCC.

Tertiary Lymphoid Structures (TLS) are lymphoid aggregates in non-lymphoid tissues and are commonly associated with chronic inflammation and cancer (9). Their presence in the tumor microenvironment (TME) provides evidence of intratumoral cooperation between the innate and adaptive arms of the immune and are often associated with improved clinical outcomes and higher responses to ICB in solid malignancies (10–13). Recently, a TLS gene signature was proven to predict response to ICB in HNSCC (14). However, their composition, size, spatial organization, and distribution within and across tumor sites in HNSCC patients treated with ICB remain understudied.

Multispectral imaging is a powerful tool for profiling the spatial architecture of the TME in solid malignancies (15,16). It allows for the spatial characterization of protein markers without altering the tissue architecture, enabling deep analysis of cell-cell spatial interaction in the TME (17). Newer scanning and image analysis platforms facilitate the use of this technology on a larger scale (18). Here, we applied high-plex imaging technology to profile the TME of pre-treatment tumor specimens of HNSCC receiving ICB.

## Materials and Methods

### Patient selection

After Institutional Review Board approval (#IRB 2022P000856), patients were retrospectively selected based on response status. Nine responders and eleven non-responders to ICB, classified via RECISTv1.1 criteria, were included. Formalin-fixed paraffin-embedded (FFPE) tissue blocks before ICB start were collected and processed for multiplex immunofluorescence (mIF) staining. Patients CTC-18, 1405_ICI-9, 1431-CTC-21, and 1408_ICI-25 had 1 replicate from the same specimen. Replicates were included in the spatial analysis (n=24). Spatial data from the slides with the biggest tissue size were included in the survival analysis and the prediction model (n=20).

### FFPE tissue specimens

Serially cut 4-µm thick sections were obtained from FFPE tumor biopsies. Human tonsils and discarded HNSCC FFPE tissue blocks were prepared for conventional immunohistochemistry (IHC), multiplex validation, and assay optimization. Before staining, all tissue slides were deparaffinized and rehydrated by serial passage through graded ethanol concentrations.

### Immunohistochemistry (IHC) validation

Single tumor sections were stained with chromogen-based IHC to validate our multiplexed immunofluorescence staining. All staining was manually performed, with antibodies against the following: Pancytokeratin AE1/AE3 (Leica/(AE3/AE3)/Mouse-IgG1, 225 mg/L), CD19 (Leica/BT51E/Mouse-IgG2B, 35 mg/L), CD56 (Leica/CD564/Mouse, 11 mg/L), CD16 (Cell Signaling/D1N9L/Rabbit IgG, 100 µg/mL), alpha Smooth Muscle Actin (SMA)(Dako/1A4/Mouse-IgG2a, kappa 71 mg/L), Ki67 (Thermo Fisher/SP6/Rabbit-IgG, 0.029 mg/ml), CD8 (Cell Signaling/D1N9L/Rabbit IgG, 28.5 mg/L), CD4 (Abcam/EPR6855/ Rabbit monoclonal-IgG, 100 µg/mL), PD1 (Abcam/EPR4877/ Rabbit monoclonal-IgG, 1.85 mg/ml), CD20 (Leica/L26/Mouse-IgG2A, kappa, 95 mg/L), LAMP3 (Thermo Fisher/PA5-84069/ Polyclonal-Rabbit-IgG, 0.1 mg/mL), CD163 (Leica/10D6/Mouse, 49 mg/L). Lot numbers are stated in Supplementary Table 1. Expression of cell markers was visualized using Vector DAB (3,3’-diaminobenzidine) Substrate kit (SK-4100). This methodology uses a diaminobenzidine reaction to detect antibody labeling, together with a hematoxylin counterstaining. A board-certified head and neck pathologist (PS) reviewed and verified positive controls for each marker.

### Spectral Library Creation

The spectral library was created according to Akoya Opal Assay Development Guide (https://www.akoyabio.com/wp-content/uploads/2020/04/Opal-Reagents_Brochure_Opal-Assay-Development-Guide.pdf). Using HNSCC tissue, single-plex slides were made for each marker with a matched fluorophore, 4’,6-diamino-2-phenylindole (DAPI), in addition to one unstained slide for autofluorescence detection. All antibodies were tested for sensitivity and specificity to determine the optimal concentrations for primary antibodies. Further optimization was done until spectral readouts for each marker were between 5-30 normalized counts (InForm, Akoya). In brief, representative areas from single-plex slides were captured at 20x magnification using our multiplexed imaging system (Vectra 3, Akoya) following spectra extraction and storing using InForm 2.5.1. The quality of the spectral library was assessed by using the R package vignettes/unmixing_quality_report.Rmd to measure unmixing quality. Raw images were spectrally unmixed using the generated libraries for later import into our data analysis platform (HALO v3.5.3577.173 Indica Labs).

### Multiplex Immunofluorescence (mIF) staining

We utilized a commercially available manual mIF staining kit (Opal 7-plex, Akoya, cat# NEL811001KT) and optimized the protocol for all markers and antibodies **(Supp. Table S1)**. Briefly, serially cut sections were deparaffinized, rehydrated, and subjected to heat-induced epitope retrieval in Tris EDTA buffer pH 9.0 (Vector Laboratories H-3301-250) or Citrate-based pH 6.0 (Vector Laboratories H-330-250) buffer (exclusively for anti-CD20 antibody) and sequential staining of each antibody was performed until completion of two panels (one in each serially cut slide) per patient (Immune and TLS panels). Nuclei were labeled using 4′,6-diamidino-2-phenylindole (DAPI, Spectral DAPI FP 1490, Perkin Elmer, 1 ug/mL). Subsequently, a coverslip with mounting media (ProLong Glass Antifade Mountant, P36980 inner, refractive index 1.52, Invitrogen) was applied and cured for at least 6 hours.

### Multispectral Imaging Platform

All slides, including those stained using IHC approaches, were imaged at 20X (NA 0.6, Vectra3, Perkin Elmer, bulb intensity set to 10%). For each tissue section, the entire tissue area was scanned. Exposure times were as follows: Immune Panel: DAPI: 35 ms, FITC: 50 ms, Cy3: 45 ms, Texas Red: 17 ms, Cy5:40. TLS panel: DAPI: 35 ms, FITC: 50 ms, Cy3: 45 ms, Texas Red: 10 ms, Cy5:40.

### Downstream mIF Image Processing

Following staining and imaging, serially cut tissue sections were registered, and a synchronous navigation tool was used to check fusion quality (HALO v3.5.3577.173 Indica Labs). Tumor-stroma classification was performed by a head and neck pathologist (PS) using Hematoxylin Eosin (H&E) stained tumor sections. Following this process, mIF-stained slides were classified using a random forest classifier trained against the pathologists’ annotations. Single-cell phenotyping was done using Indica Labs – HighPlex FL v4.1.3 module **(Supp. Table S2)**. A detectable nucleus was mandatory to phenotype each cell. The same thresholds were applied to all the slides, and spatial analysis was performed based on average values.

### Tertiary Lymphoid Structures quantification

Tertiary Lymphoid Structures were defined as aggregates of lymphoid cells with histologic features resembling follicles in lymphoid tissue and were identified on a serially cut H&E slide by a head and neck pathologist (PS). TLS were identified on serially cut slides stained with mIF panels using random forest classifiers as described above. At single-cell mIF phenotyping, we included TLS with more than fifty B cells (CD20+) and more than five dendritic cells (LAMP3+). The number of TLS was normalized by the area analyzed (TLS/mm2). The average distance to the tumor area was calculated for all patients. The peritumoral area was defined as <100 µm from the tumor area (19)

### Survival analysis

The Kaplan-Meier analysis was applied using the Log-rank test to assess the association between the spatial location of TLS and both OS and PFS. Hazard ratios (HR) and 95% CI were computed. Given the sample size and the potential variability in the data, we employed a bootstrapping approach to obtain more robust estimates of the hazard ratios and their associated confidence intervals. Specifically, we performed 1,000 bootstrap resampling iterations (R = 1000), where the original dataset was repeatedly resampled with replacement. For each bootstrap replicate, the Cox model was refit, and the hazard ratio was recalculated. All values were considered statistically significant if a < 0.05. Survival analyses were performed with the survival (v3.5.5) and survminer (v0.4.9) packages using R Statistical Software (v4.3.1, R Core Team 2023).

### Machine learning model

To predict the response to ICB using spatial metrics, including TLS features, we evaluated several multiclass classifiers, including Logistic Regression, Decision Tree, ExtraTree, Random Forest, and Gradient Boosting. We used an initial set of features consisting of: 1. The density of CD8+PD1+Ki67- in the tumor/density of CD8+PD1+Ki67- in the stroma (CD8 ratio); 2. The density of CD20+ cells/density of CD163+ cells (CD20 ratio), Combined Positive Score; 3. The average size of TLS (bigger or smaller than the mean size in the whole cohort); 4. TLS density (# of TLS/tissue size) via mIF. We also included spatial metrics related to immune cell interactions.

Missing values were imputed using *SimpleImputer* from scikit-learn. Continuous features were then standardized using StandardScaler. We trained the models using a 75:25 training/validation split, and a search approach was used to test combinations of features to select the set that resulted in the highest accuracy. To understand the contribution of each variable, we visualized permutation importance. We assessed the robustness of the results through 5-fold cross-validation. Finally, we recorded model accuracy, precision, recall, F1-score, and area under the curve (AUC).

In addition to classifier approaches, we validated findings using a convolutional neural network to predict ICB response directly from whole slide images. For this analysis, we preprocessed the images by resizing them to 224x224 pixels and normalized pixel values to the range [0, 1]. We augmented the image dataset using random rotations, shifts, flips, and zoom to improve model generalization. We adopted a transfer learning approach using a pre-trained ResNet50 model as a feature extractor. The base model was modified by removing top layers and adding a custom head consisting of a flattening layer, dense layers, and dropout for regularization. The last 10 layers of the base model were fine-tuned on our dataset. The model was trained using binary cross-entropy loss and the Adam optimizer. We implemented early stopping to prevent overfitting and evaluated model performance using 1,000 bootstrap resamples. This approach allowed us to assess whether whole slide images, in combination with extracted features, could predict ICB response. To understand features driving model predictions, we used Gradient-weighted Class Activation Mapping (Grad-CAM) to visualize regions of the whole slide images that were most influential in the model’s decision-making. Analyses were performed using Python v3.11.4, Jupyter notebook v6.5.4, scikit-learn v1.3.0, and TensorFlow v2.15.0.

## Results

### Patient characteristics

Patient clinicopathological features are stated in **Table 1**. The average age at the start of ICB was 62 years old (range 50-81). The oral cavity was the most common primary site. High (≥20) and moderate (1–19) CPS were observed in most of the cohort. CPS did not differ between responders and non-responders (*unpaired t-test, p= 0.3649*) **(Supp. Figure 1. A)**. Fourteen (70%) patients received pembrolizumab as monotherapy, and six (30%) patients received ICB plus chemotherapy.

**Table 1.** Clinical characteristics of the cohort.

| Total cases | n=20 | Responders | Non-responders | Difference among groups (t-test) |
| --- | --- | --- | --- | --- |
| Sex |  |  |  | 0.50 |
| Male | 16 | 8 | 8 |  |
| Female | 4 | 1 | 3 |  |
| Age at ICB start, median (range, year) | (62, 50-81) |  |  | 0.80 |
| <65 | 14 | 5 | 9 |  |
| ≥65 | 6 | 4 | 2 |  |
| Anatomic site |  |  |  | 0.60 |
| Oral cavity | 11 | 4 | 7 |  |
| Oropharynx | 6 | 3 | 3 |  |
| Nasopharynx | 2 | 1 | 1 |  |
| Larynx | 1 | 1 | 0 |  |
| Ethnicity |  |  |  | 0.47 |
| Hispanic | 1 | 1 | 0 |  |
| White non-Hispanic | 16 | 6 | 10 |  |
| Asian | 2 | 0 | 2 |  |
| Non available | 1 | 1 | 0 |  |
| ECOG performance status |  |  |  | 0.59 |
| 0-1 | 15 | 6 | 9 |  |
| 2 | 2 | 1 | 1 |  |
| 3 | 0 | 0 | 0 |  |
| 4 | 0 | 0 | 0 |  |
| Non available | 3 | 2 | 1 |  |
| Treatment |  |  |  | 0.69 |
| Pembrolizumab | 13 | 5 | 8 |  |
| Nivolumab | 1 | 0 | 1 |  |
| Pembrolizumab + chemotherapy | 6 | 4 | 2 |  |
| Tobacco use |  |  |  |  |
| Never | 9 | 4 | 5 | 0.18 |
| Former | 9 | 4 | 5 |  |
| Non available | 2 | 1 | 1 |  |
| Alcohol consumption |  |  |  | 0.69 |
| Current | 13 | 6 | 7 |  |
| Former | 2 | 2 | 0 |  |
| Never | 5 | 1 | 4 |  |
| TNM stage 1 |  |  |  | 0.66 |
| I | 0 | 0 | 0 |  |
| II | 2 | 2 | 0 |  |
| III | 5 | 2 | 3 |  |
| IV-IVC | 13 | 5 | 8 |  |
| Viral status |  |  |  | 0.77 |
| HPV | 5 | 4 | 1 |  |
| Negative | 7 | 3 | 4 |  |
| Non available | 8 | 2 | 6 |  |
| CPS |  |  |  | 0.631 |
| <1 | 3 | 1 | 1 |  |
| 1 - 19 | 8 | 5 | 4 |  |
| ≥20 | 9 | 3 | 6 |  |
| Best Overall Response to ICB (RECIST v1.1) |  |  |  | 1 |
| PD | 11 | 0 | 11 |  |
| SD | 2 | 2 | 0 |  |
| PR | 4 | 4 | 0 |  |
| CR | 3 | 3 | 0 |  |
| Number of infusions |  |  |  | 0.86 |
| ≤5 | 9 | 1 | 8 |  |
| 5-20 | 8 | 6 | 2 |  |
| ≥21 | 3 | 2 | 1 |  |
| Number of prior systemic therapies |  |  |  | 1 |
| 1 | 5 | 4 | 1 |  |
| 2 | 4 | 2 | 2 |  |
| 3 | 3 | 1 | 2 |  |
| 4 | 3 | 2 | 1 |  |
| 5 | 3 | 0 | 3 |  |

PD: Progressive disease, SD: stable disease, PR: partial response, CR: complete response

### Spatial Distribution of Cells in the Tumor Microenvironment

Multispectral profiling of individual cells within tumor tissues allowed us to characterize the distribution of 12 proteins within the tumor microenvironment (TME) **(Figure 1. A, B)**. We found no difference in the area classified as tumor or stroma based on response to ICB (*unpaired t-test p=0.6470, p=0.6108,* respectively) **(Figure 1. C)**. Cell distribution varied across compartments in the TME. In the area classified as tumor, the proliferation marker (Ki67+) contributed to 35% and 36% of positive cells for responders and non-responders, respectively. Interestingly, immature B cells (CD19+ cells) contributed 10% to responders and 3% to non-responders. The stromal area was characterized by the presence of fibroblast (α-SMA) (26% in responders and 20% in non-responders), followed by natural killer cells (CD16+) (15%) for responders and CK+ cells for non-responders (19%) **(Figure 1. C)**.

**Figure 1.**
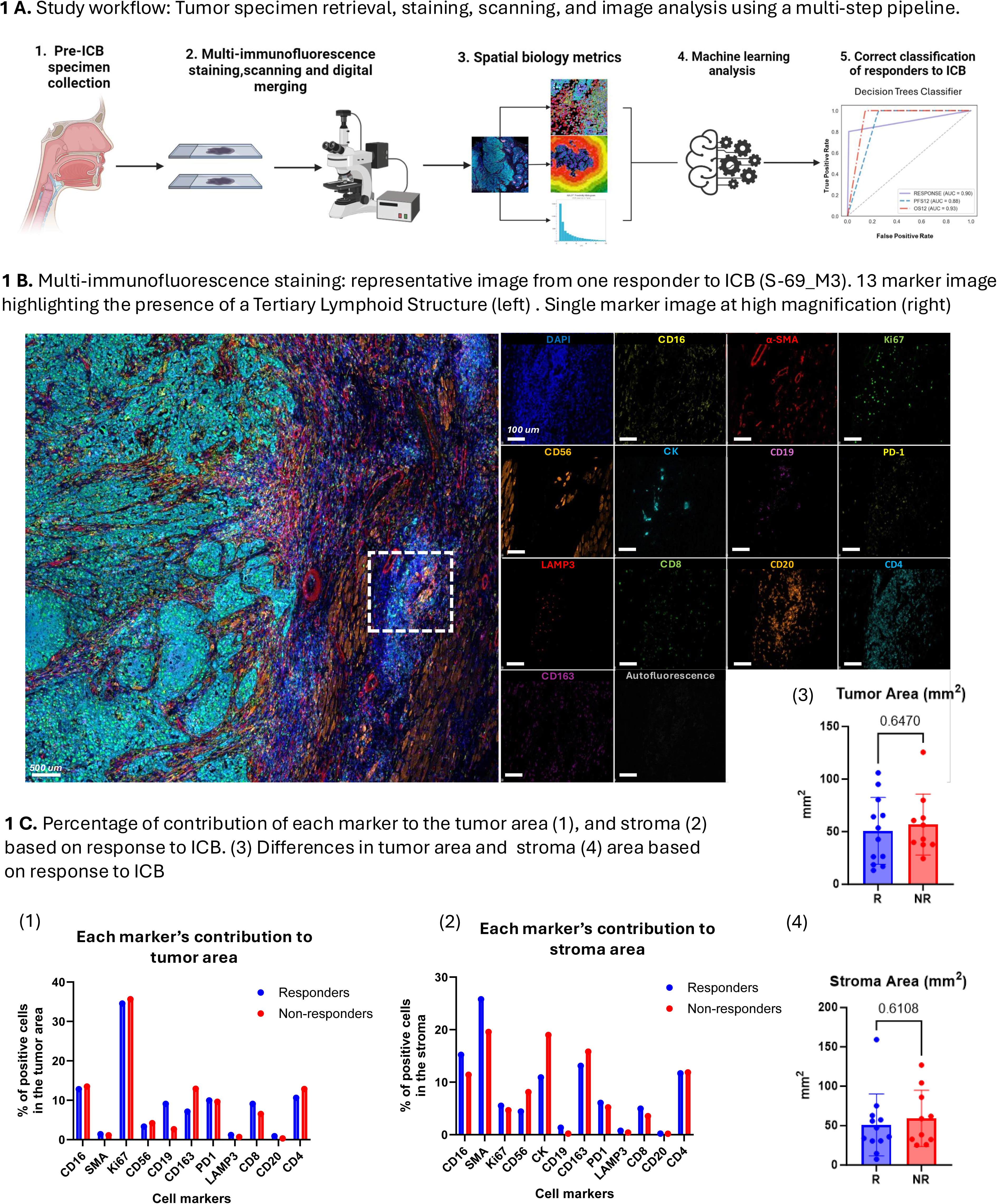
Study workflow and characterization of the tumor-immune microenvironment in HNSCC. **1 A.** Overview of study workflow, tumor specimen retrieval, staining, scanning and image analysis using a multi-step pipeline including machine learning algorithms. Sequentially cut slides were stained with different staining panels, scanned, and fused using our image analysis platform (HALO, Indica Labs). **1 B.** Multi-immunofluorescence staining was performed; a representative image from patient S-69_M3 is displayed here. All markers are shown (left), and a single marker view (right) is shown. **1 C.** Each marker’s contribution percentage to the tumor area (1) and stroma (2) is shown based on response to ICB. Additionally, differences between the tumor and stroma areas based on response to ICB are shown (3,4).

### Spatial analysis identifies increased B cell density in responding patients

We found no significant difference between the average distance of multiple cell phenotypes to CK+ tumor cells or tumor area based on response status **(Supp. Figure 1, 2; Supp. Table 3)**. However, the spatial architecture within immune populations revealed interesting trends in the organization of dendritic cells (LAMP3+) in the TME. A trend toward an increased number of exhausted T cells (CD8+PD1+Ki67-) within 20 um of dendritic cells (LAMP3+) was found in responders compared to non-responders (*unpaired t-test p= 0.0576)* **(Figure 2. A, B)**. Additionally, a trend toward higher density of proliferative dendritic cells (LAMP3+PD1-Ki67+) was found in responders compared to non-responders (*unpaired t-test p= 0.22*) **(Figure 2. C)**. Interestingly, a significantly higher density of B cells (CD20+) was observed in responders compared to non-responders (u*npaired t-test p= 0.02*) **(Figure 2. D, E)**. No other difference in densities was found among groups **(Supp. Figure 3; Supp. Table 4)**. The interplay of B cells with other cells is critical for the development of TLS, leading us to investigate these structures in the TME.

**Figure 2.**
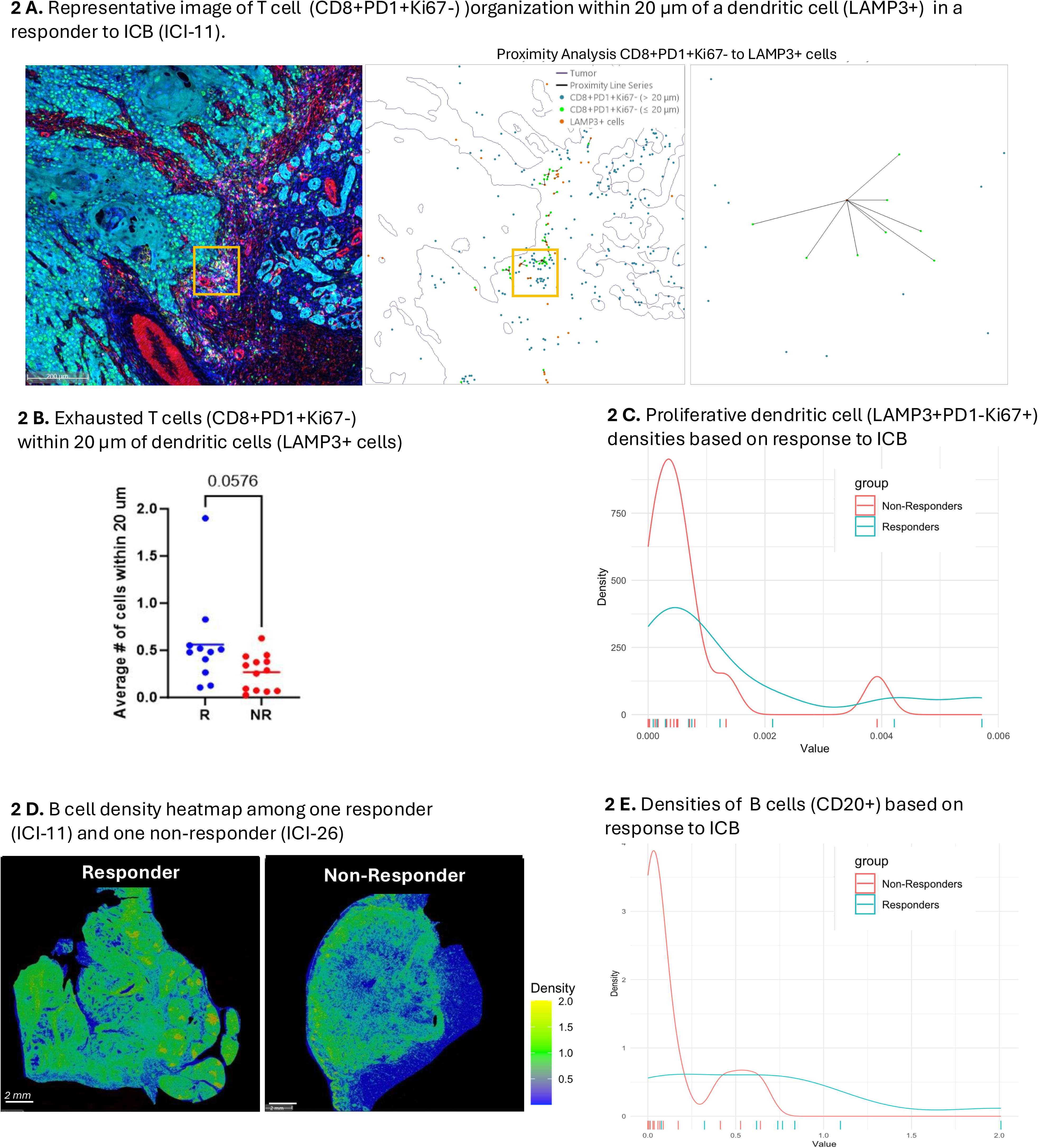
Spatial analysis of HNSCC TME reveals preferential organization of antigen-presenting cells. **2 A.** A representative image of T cell (CD8+PD1+Ki67-) organization within 20 µm of a dendritic cell (LAMP3+) in a responder to ICB (ICI-11). A proximity analysis between T cells and dendritic cells is shown (right). **2 B.** Average number of T exhausted T cells (CD8+PD1+Ki67-) within 20 µm of dendritic cells (LAMP3+) is shown. **2 C.** A rug plot displays the proliferative dendritic cell (LAMP3+PD1-Ki67+) densities based on response to ICB. **2 D.** B cell density heatmap among one responder (ICI-11) and one non-responder (ICI-26). **2 D.** A rug plot shows the densities of B cells (CD20+) based on response to ICB.

### The organization of TLS within the TME impacts survival

The presence of TLS was assessed first on H&E slides by a head and neck pathologist (PS), who provided a raw number of TLS per slide. Using their annotations and setting a threshold for a minimum of 5 dendritic cells (LAMP3+) and 50 B cells (CD20+) per structure, the presence of TLS was subsequently assessed via mIF **(Figure 3. A)**. The detection rate of at least one TLS on the H&E slide by the pathologist was 50% (12/24) compared to 67% via mIF (16/24) for sequentially sectioned mIF slides. Importantly, a higher number of TLS was detected via mIF for all samples except one (1408_ICI-25), for which the amount (one TLS) was the same for both approaches. For the slides with replicates, a variation of <1 TLS was observed via pathologist assessment, but a variation of 4.7 (0-9) unique TLS was observed among replicates using mIF **(Supp. Table 5)**.

**Figure 3.**
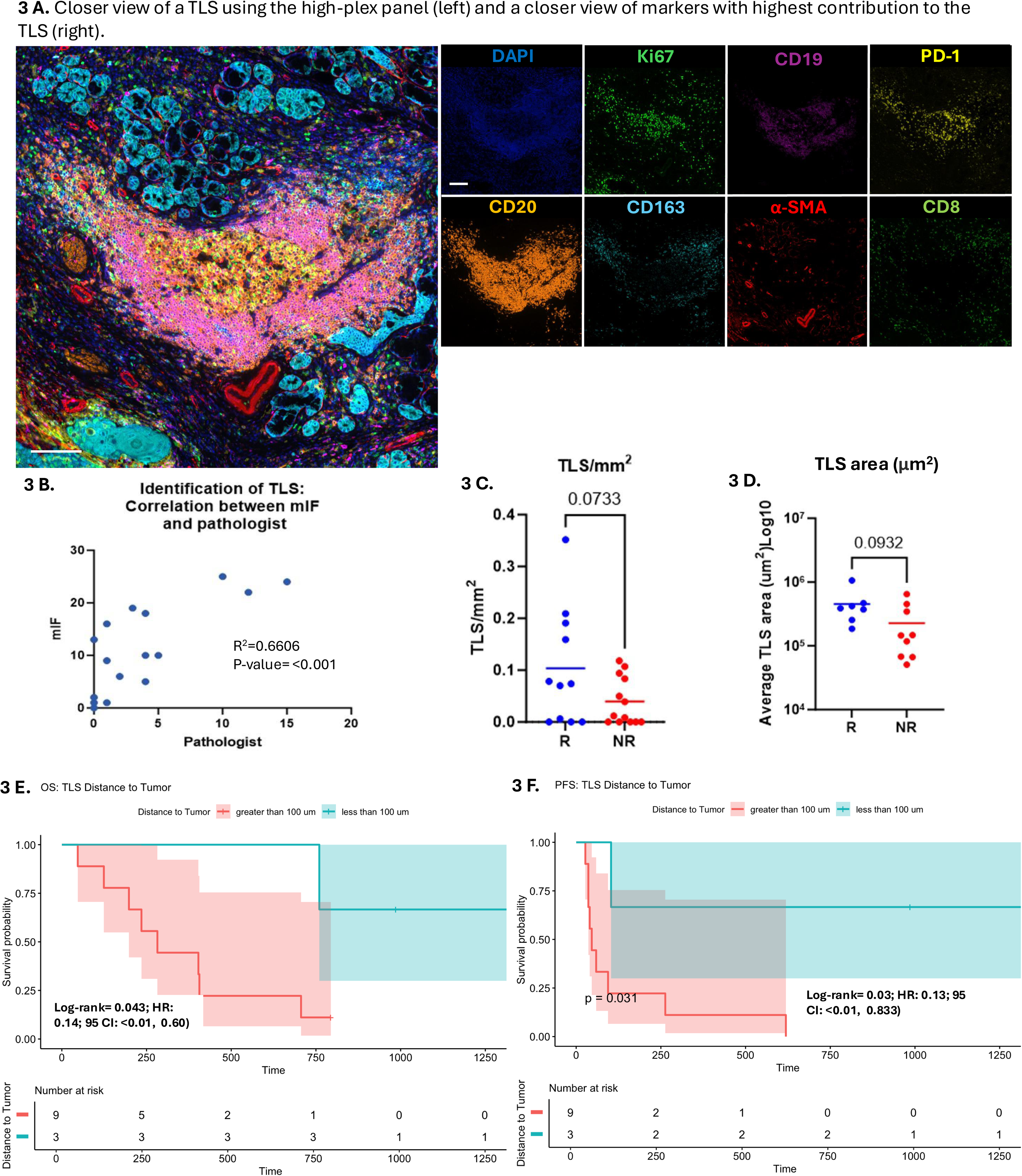
The spatial organization of tertiary lymphoid structures impacts survival. **3 A.** A closer view of a TLS is displayed using high-plex immunofluorescence (left) and the contribution of each marker to the TLS (right). **3 B.** Correlation between mIF and pathologist in identifying TLS in serially cut slides. **3 C.** Number of TLS normalized by the tissue size. **3 D.** Average TLS area in responders compared to non-responders. **3 E, F.** Overall and progression-free survival based on the average distance of TLS to the tumor area.

The identification of TLS using mIF was positively correlated with the assessment done by the pathologist on serially cut H&E sections (*R^2^= 0.66, p-value= <0.0001*) **(Figure 3.B)**. When normalized by tissue size, responders tended to have higher TLS/mm^2^ compared to non-responders (*unpaired t-test, p-value= 0.0733*) **(Figure 3. C) (Supp. Table 5)**. These structures were constituted mainly by CD4+ T cells (21%, range 14-40), followed by CD20+ B cells (17%, range 8-36) **(Supp. Fig. 4. A; Supp. Table 6)**. No difference was found between the distance of TLS to tumor area among groups **(Supp. Figure 4. B)**. Importantly, responders tended to have a higher TLS area on average compared to non-responders (*unpaired t-test; p-value= 0.0932*) **(Figure 3. D),** but there was no difference in overall survival (OS) or progression-free survival (PFS) between groups based on the size of TLS **(Supp. Figure 5. A-B)**. Among the 12 patients with TLS, those whose average TLS distance to the tumor was within 100 µm had significantly better outcomes. Specifically, the median OS was 33.5 months for patients with TLS within 100 µm of the tumor area, compared to 11.6 months for those with a distance greater than 100 µm (Log-rank p=0.05). Similarly, the median PFS was 26.3 months for patients with TLS within 100 µm of the tumor area versus 4.4 months for those in which the average TLS-tumor distance was greater than 100 µm (Log-rank test p=0.03) **(Figure 3. E-F).**

### A multivariable machine learning model allows for the prediction of response and survival

The response to ICB involves dynamic interactions between multiple components in the immune system and how those are organized in the TME. Therefore, we decided to investigate the impact of immune cell ratios on HNSCC clinical outcomes. No difference was found between the ratio of exhausted T cells in the tumor to the stromal area (CD8+PD1+Ki67- density in the tumor area/CD8+PD1+Ki67- density in the stroma) and clinical outcomes **(Suppl. Fig 6. A-C)**. Calculating the ratio of CD20/CD163 densities, we observed a trend towards short OS (HR 0.43, 95% CI: 0.05-3.39; p-value= 0.4) and PFS (HR 0.3, 95% CI: 0.04-2.83; p-value= 0.3) for patients whose ratio was below the average for the cohort (<2.57), underscoring the potential pro-tumoral potential of CD163+ cells **(Suppl. Fig 7. A)**.

Spatial metrics were associated with response to ICB therapy. Natural killer cells (CD16+Ki67-) trended to be closer to proliferative dendritic cells (LAMP3+Ki67+) in responders compared to non-responders (p= 0.197) **(Figure 4. A, B),** and this average distance trended to be negatively correlated with response to ICB (p=0.098), with a higher probability of response for patients with shorter distances between these two cell populations. **(Figure 4. C)**.

**Figure 4.**
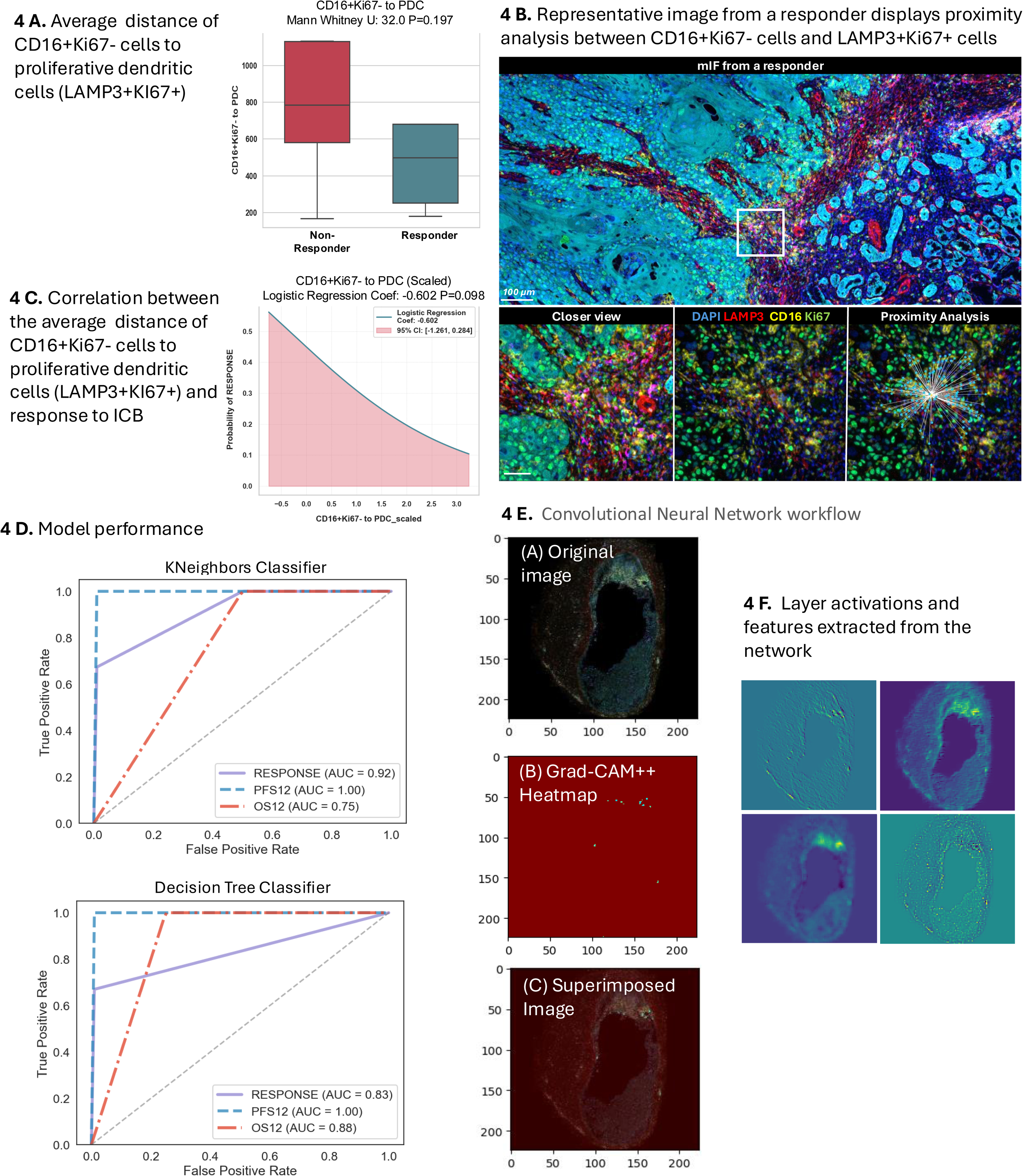
A multivariable machine learning model allows for the prediction of response to ICB and survival in HNSCC. **4 A.** Comparison between the average distance of natural killers (CD16+Ki67-) to proliferative dendritic cells (LAMP3+Ki67+) based on the response to ICB. **4 B.** Representative image from the analysis in **4 A.,4 C.** Correlation between the average distance of natural killers (CD16+Ki67-) to proliferative dendritic cells (LAMP3+Ki67+) and response to ICB. **4 D.** Model performance: Area under the curve (AUC) for the models used to predict response to ICB. **4 E.** Convolutional Neural Network workflow. (A) Original Image: Cross-sectional medical image showing tissue structure. (B) Grad-CAM++ Heatmap: Visualization of regions critical for model classification. Brighter areas indicate higher importance, and darker areas (blue/green) indicate less importance. (C) Superimposed Image: Original image overlaid with Grad-CAM++ heatmap, highlighting model-focused areas within tissue context. **4 F**. Layer Activations: Four feature maps from the initial convolutional layer of the ResNet model. Each panel represents a distinct activation channel, revealing different low-level features extracted by the network.

Subsequently, a multivariate model was used to predict the response of each patient based on the spatial metrics. The best-performing model was the K-Nearest Neighbors classifier, which achieved an accuracy of 0.80 (AUC 0.92) in predicting response, 1.00 (AUC 1.00) in predicting 12-month progression-free survival (PFS), and 0.80 (AUC 0.75) in predicting 12-month overall survival (OS) **(Figure 4. D)**. Feature importance analysis demonstrated that the density of TLS was the leading predictor consistently across all model architectures **(Supp Figure 8)**. Other important features in the models included the ratio of CD8+PD1+ in the tumor/stroma, the CD20+/CD163+ ratio, and the LAMP3 stromal density **(Supp Figure 8)**. These findings support that TLS density and spatial organization of immune components play an important role in determining the immune response and clinical outcomes in HNSCC patients treated with ICB.

A CNN was then trained to predict response directly from unannotated whole slide images using the dataset of twenty patients. When combined with the best-performing K-Nearest Neighbor model in an ensemble approach, the model achieved promising results in a small test set of five patients: an accuracy of 0.81 (95% CI: 0.40 - 1.00) in predicting response, 1.00 (95% CI: 0.40 - 1.00) in predicting one-year progression-free survival (PFS), and 0.86 (95% CI: 0.57 - 1.00) for one-year overall survival. To provide additional insight, a saliency map was generated to visualize and interpret the features driving the model’s predictions. This map offered insight into the image characteristics, including Tertiary Lymphoid Structure-related features, that appeared most influential in determining patient outcomes (**Figure 4. E, F**).

## Discussion

Here, we applied multi-spectral imaging methods to quantify twelve markers in pre-ICB HNSCC tumor specimens, identified critical spatial signatures uniquely found in patients with effective responses to ICB, and differentially profile patients with a higher likelihood of response to ICB. We found improved progression-free survival in patients whose average TLS epicenter-tumor edge distance was within 100 µm, highlighting the importance of spatial localization in biomarker studies. Additionally, with a machine learning model consisting of spatial biology metrics, together with raw image features, we were able to predict response and one-year overall survival with 81% and 86% accuracy, respectively. Importantly, TLS density was the leading predictor across models. CPS provides a simple, single-marker view, whereas the multivariate model captures the complex relationship between diverse immune populations that are important for response. The spatial metrics outperformed CPS, highlighting how incorporating spatial biology can improve predictive models beyond conventional biomarkers.

Although CPS is used daily in the clinic, its effectiveness as a predictive biomarker for ICB response is limited. Patients with low or negative CPS can still respond to ICB, while those with high CPS may not always experience a positive response (4,20–23) Additionally, using a single marker to stratify a patient’s likelihood of response has limitations. First, whole tissue scoring without discerning between tumor or stroma ignores the impact of cell localization within the TME. Second, PD-L1 is expressed in multiple cell phenotypes with various roles in response to ICB. Therefore, PD-L1 positivity alone might not be enough to profile the TME, particularly considering its variation across time (24). In our study, we seek to address these limitations by using multi-step high-plex imaging technology.

Dendritic cells are antigen-presenting cells and are responsible for priming CD4 and CD8 T cells (25). Physical interactions between subsets of T cells and dendritic cells have been shown to control antitumor immunity (26). Here, we saw a trend toward increased average number of exhausted T cells (CD8+PD1+Ki67-) within 20 µm of dendritic cells (LAMP3+) in responders compared to non-responders. This spatial localization might facilitate T cell priming by dendritic cells in the context of ICB, resulting in an improved antitumor response. LAMP3+ dendritic cells have been shown to interact with neoantigen-reactive exhausted CD8 T cells and CD4 T regs in cervical cancer, where LAMP3 was associated with an immunosuppressive state, leading to tumor escape and disease progression (27). In contrast to our results, no statistical significance was found in the density of dendritic cells (LAMP3+) among responders compared to non-responders. Our limited sample size may have biased these results and future investigations with more patients will be necessary to fully support these findings.

B cells produce antibodies specific to tumor-associated epitopes that can boost anti-tumor T cell response and can act as antigen-presenting cells (28). Tumor-infiltrating B cells have been correlated with improved outcomes in multiple solid malignancies (29–32) and have been shown to play a critical role in response to ICB (10,11,33,34). Interestingly, higher densities of CD20+ B cells and their physical interaction with CD8 T cells were associated with improved prognosis in treatment-naive HPV+OPSCC patients despite analyzing only ten representative low-powered visual fields per patient (35). Our results are aligned with prior studies and support the anti-tumor role of B cells in the TME (36,37).

The ratio of peri-tumoral CD8/FOXP3 densities is useful in detecting patients who are more likely to respond to ICB in multiple solid malignancies (38–40). Here we found no difference in survival outcomes for patients based on their tumor/stroma ratio of exhausted CD8 T cell densities **(Supp. Fig 6)**. T cell exhaustion is a gradual process, with terminally exhausted T cells being the less functional on the activation-exhaustion spectrum (41), a particular phenotype that could have been missed by our 12 markers approach. A greater characterization of exhaustion proteins could help further classify T cells and their response to ICB. Newer technologies with increased capacity for the detection of spatial transcriptomic/proteomic data (41,42) are paving the way for personalized medicine and could provide greater insights into the T cell – ICB interaction.

TLS are lymphoid structures localized within the TME, formed through interactions between the innate and adaptive immune systems (9). They are characterized by a germinal center composed of specialized immune cell types (42) and are created through the local accumulation of *CXCL13*, *RANKL*, and interleukin-7 (43). TLS have been well-established as positive prognostic factors in ICB (11,44). In HNSCC, the presence of TLS has been associated with favorable outcomes and improved responses to ICB regardless of HPV status (14,45). In our study, the presence or absence of TLS was not associated with survival outcomes **(Supp. Fig. 9)**. However, among patients with TLS, those with TLS located within 100 µm of the tumor area showed significantly improved OS and PFS **(Figure 3)**. These findings align with prior studies, which emphasize the importance of TLS spatial location over their mere presence (46). However, reported peritumoral regions for TLS vary, ranging from 1000 µm to 5000 µm from the tumor boundary (47). Larger studies are needed to reach a consensus on the predictive value of TLS spatial organization. Through multivariate modeling, we identified TLS density as the leading predictor of ICB response, outperforming conventional biomarkers such as CPS. Notably, TLS can be measured in H&E slides, and recent research has demonstrated the feasibility of automated computational workflows for measuring TLS density in lung adenocarcinoma (48,49). This highlights the promise of TLS as a relatively easy-to-detect biomarker with previously underestimated clinical significance.

Our study has limitations, including a small sample size and its retrospective design. As such, these findings should be viewed as preliminary and indicative of the potential of this approach rather than definitive evidence of its efficacy. Additionally, 30% (6/20) of our cohort received ICB in combination with chemotherapy. In these cases, the response observed in the four patients may not have been immune-driven. Nevertheless, our results suggest that TLS holds promise as an easily identifiable biomarker that could be incorporated into the prognostic workup for HNSCC. However, larger studies are needed to further assess the impact of TLS on the response to ICB.

## Conclusion

We found that TLS density strongly predicts response to ICB therapy in HNSCC, outperforming the current standard biomarker PD-L1-based CPS. Our multivariate modeling successfully integrated multiple spatial metrics related to the organization and composition of immune cells within the tumor microenvironment. These findings show the importance of immune spatial patterning in driving anti-tumor immunity. They provide a rationale for developing therapies promoting favorable TLS formation and composition. Moreover, the results support the incorporation of advanced multiplexed imaging techniques that can measure the spatial organization of the tumor microenvironment for future ICB predictive biomarker development.

## Author contributions

S.L.S., D.L. F., and M.S.F. conceptualized the investigation and provided supervision. D.R.T, S.H., and E.L. performed sample processing, staining, and scanning. D.A.R.T., M.E.B, and P.S. performed formal analysis, including visualization. R.M., T.R., M.P., J.C.P., and L.J.W. assisted with study design and sample acquisition. D.A.R.T. wrote the original draft, and all authors performed critical reviews and revisions.

## Supplementary Figure Legends

**Supplementary Figure 1.** A. CPS score based on response. B. Average distance of each marker to CK+ cell. C. Average distance of key markers to proliferative dendritic cells (LAMP3).

**Supplementary Figure 2. A.** Average distance to tumor/stroma interface. B. Distance to tumor area.

**Supplementary Figure 3.** Densities among responders and non-responders.

**Supplementary Figure 4.** TLS characterization. A. Contribution of each marker to TLS. B. Average distance of TLS to tumor area.

**Supplementary Figure 5.** Survival outcomes based on average TLS size

**Supplementary Figure 6.** Tumor/stroma ratio of densities of exhausted T cells (CD8+PD1+Ki67-) and survival outcomes are based on higher or lower than the mean value for the cohort. A. Overall survival for the ratio of exhausted T cells in the tumor/stroma. B. Progression-free survival for the ratio of exhausted T cells in the tumor/stroma. C. Representative mIF image showing a strong presence of exhausted T cells in the tumor.

**Supplementary Figure 7.** The ratio of CD20/CD163 densities for the whole slide and survival outcomes are based on higher or lower than the mean value for the cohort. A. Overall survival and B. progression-free survival. C. Representative image of CD20 and CD163 cells in the tumor microenvironment of a responder to ICB.

**Supplementary Figure 8.** Feature Importance for Multiclass Classifiers. TLS/µm (mIF): density of TLS via mIF; TLS area (mm^2^): TLS density using raw counts assessed by the pathologist. CD8+PD1+Ki67-_20µm_DC: average quantity of exhausted T cells in 20 µm radius distance of dendritic cells (LAMP3). CD8RATIO: ratio of CD8+PD1+Ki67+ in tumor/stroma. IA_CD8+PD1-Ki67+: Average distance of proliferative CD8+ T cells to the tumor area. CD16+Ki67- to PDC: Average distance of CD16+Ki67- cells to proliferative dendritic cells (LAMP3+Ki67+). TLS_size: raw average size of TLS. LAMP3STROMA: Density of dendritic cells (LAMP3+) in stroma area. CPS: Combined positive score. CD20RATIO: Ratio of CD20/CD163 densities in the whole tissues section.

**Supplementary Figure 9.** Presence or absence of TLS. A. Overall survival. B. Progression-free survival.

## Data Availability

All data produced in the present study are available upon reasonable request to the authors

