## Supplementary Figure 1 for "Immune Cell Densities Predict Response to Immune Checkpoint-Blockade in Head and Neck Cancer"

**Supplementary Figure 1. A.** CPS score based on response. B. Average distance of each marker to CK+ cell. C. Average distance of key markers to proliferative dendritic cells (LAMP3)

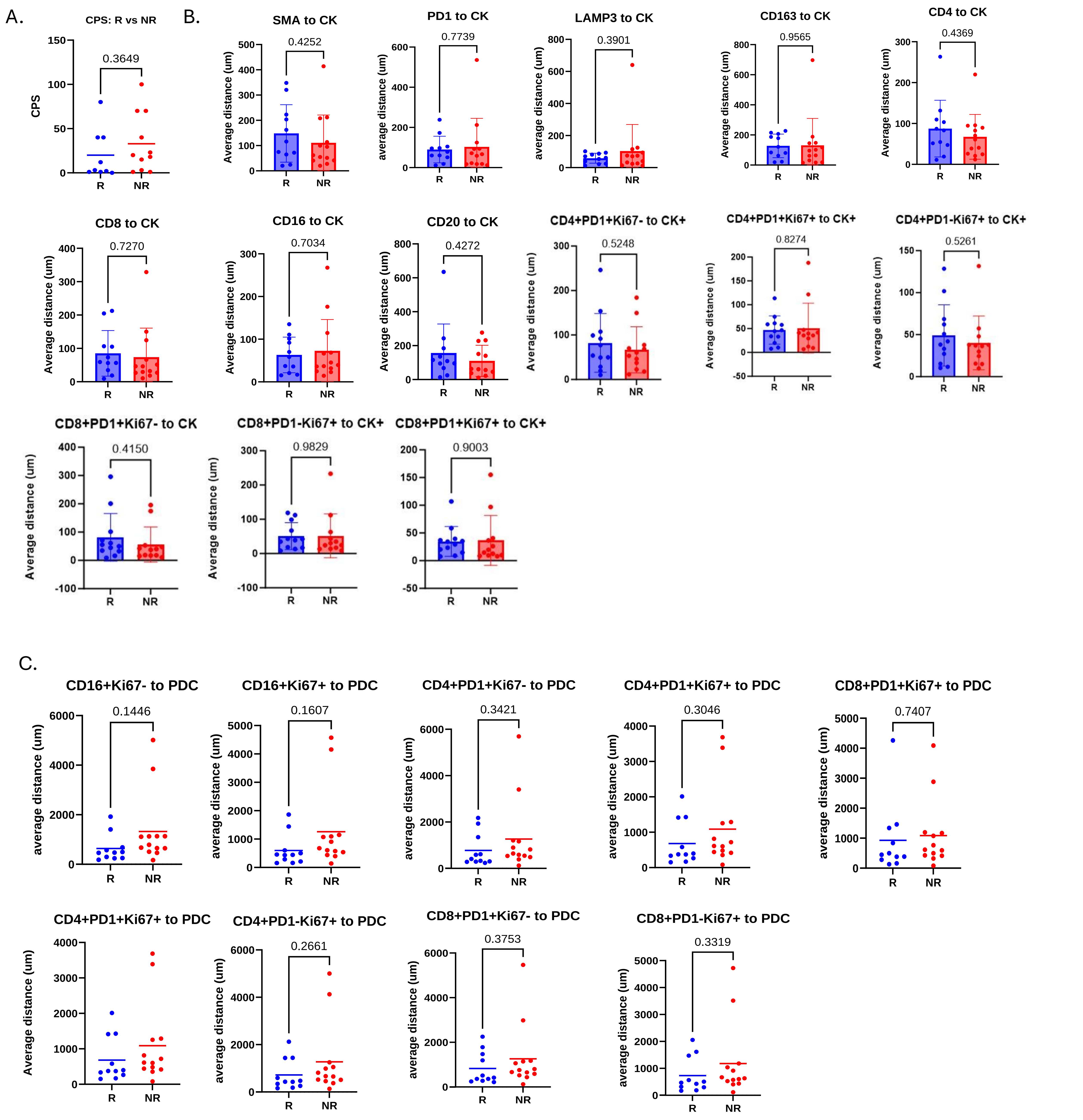
