## Supplementary figures and images for "Immune Cell Densities Predict Response to Immune Checkpoint-Blockade in Head and Neck Cancer"

### Supplementary Figure 2

**Supplementary Figure 2.** A. Average distance to tumor/stroma interface

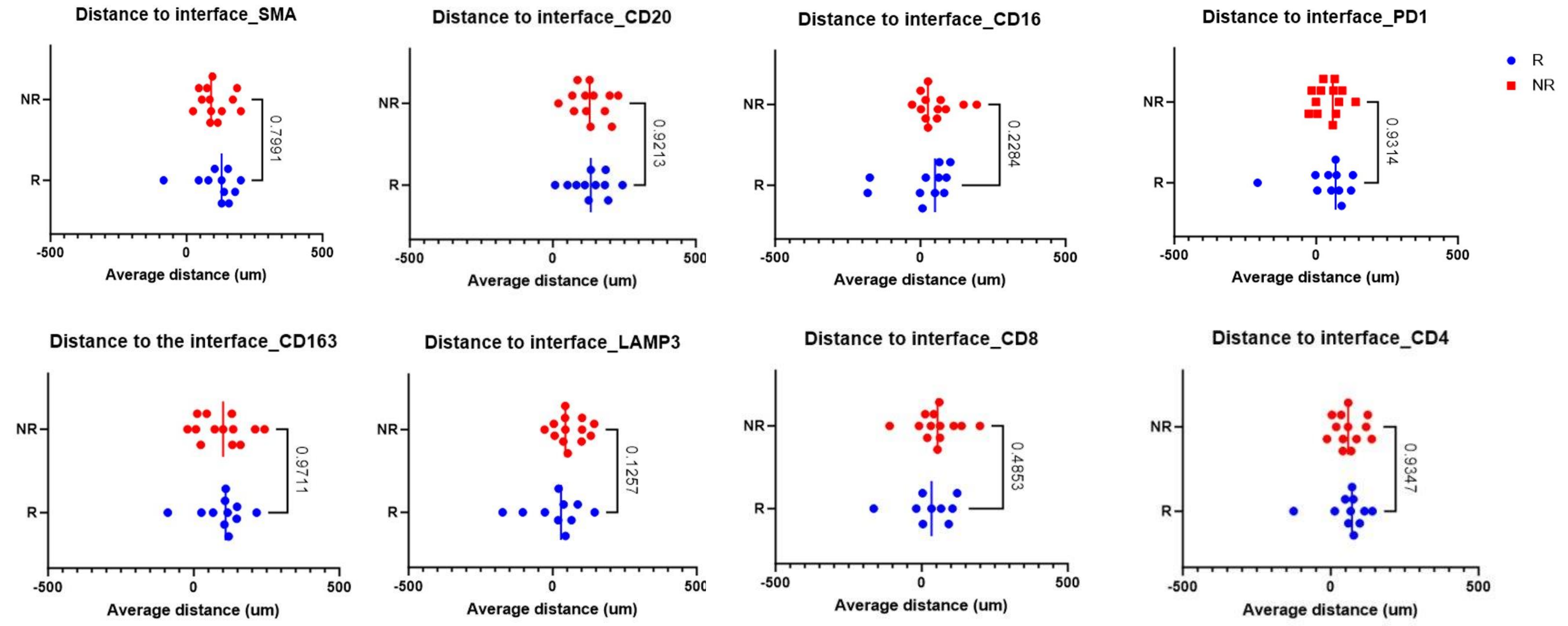


B. Distance to tumor area

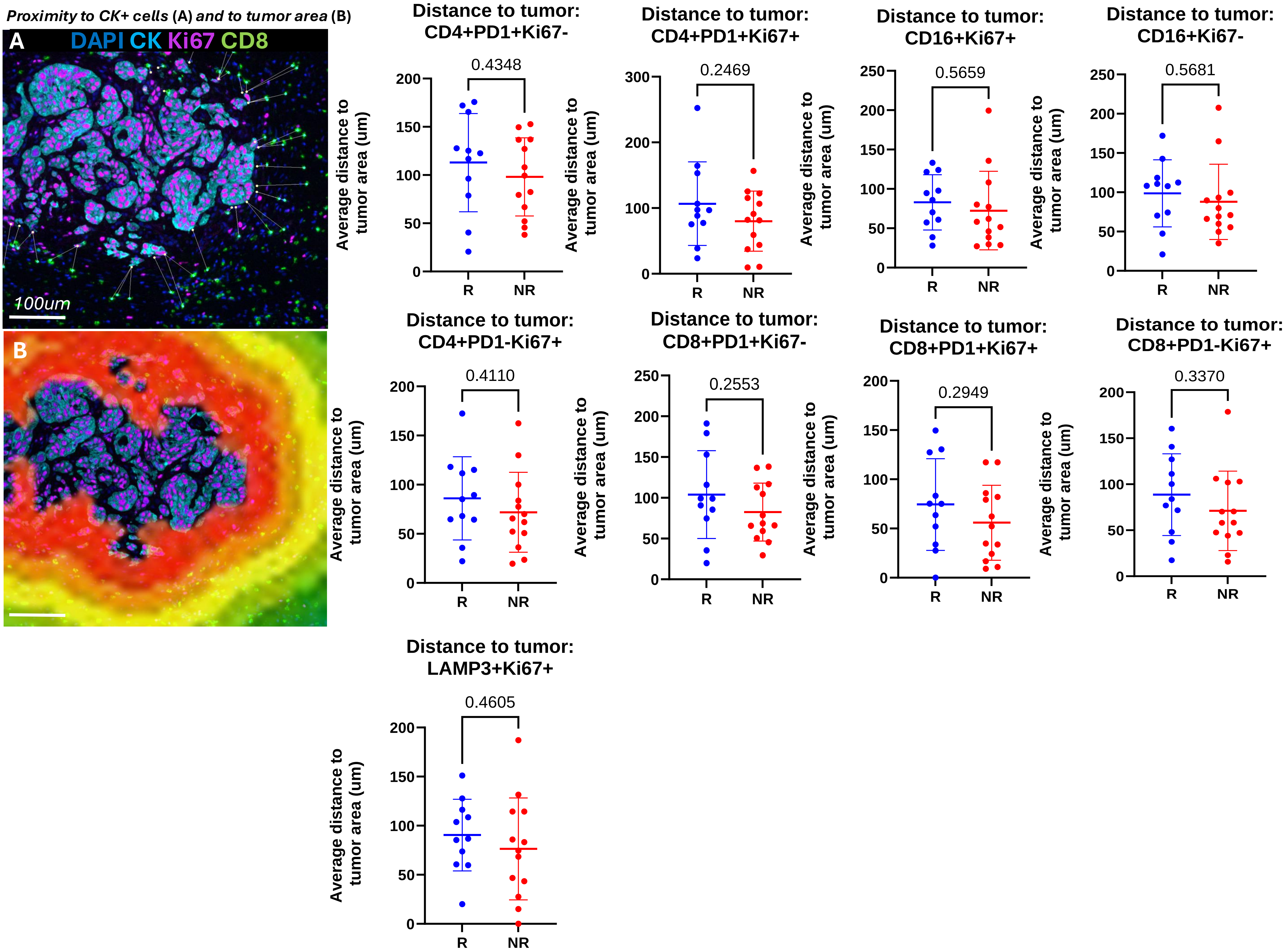
