## Supplementary Figure 3, 4 and 5 for "Immune Cell Densities Predict Response to Immune Checkpoint-Blockade in Head and Neck Cancer"

**Supplementary Figure 3.** Densities among responders and non-responders

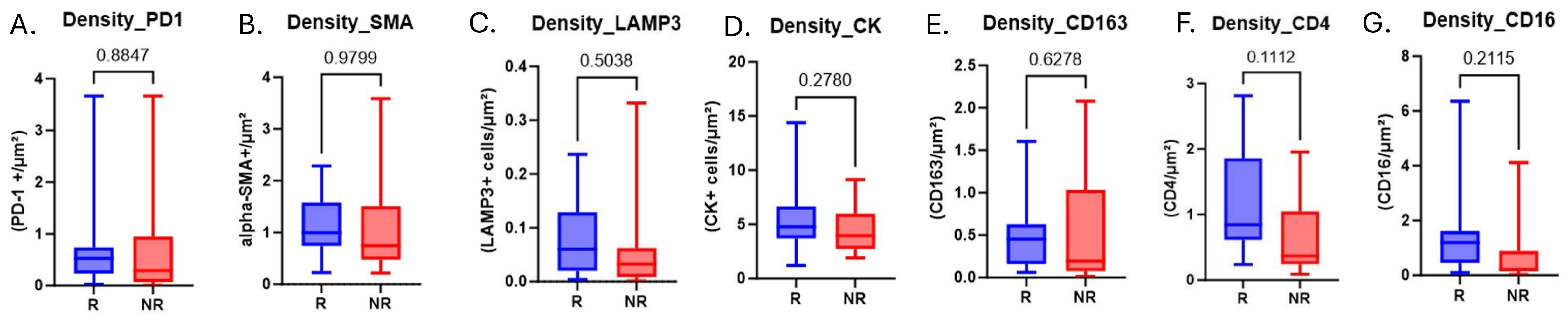


**Supplementary Figure 4.** TLS characterization

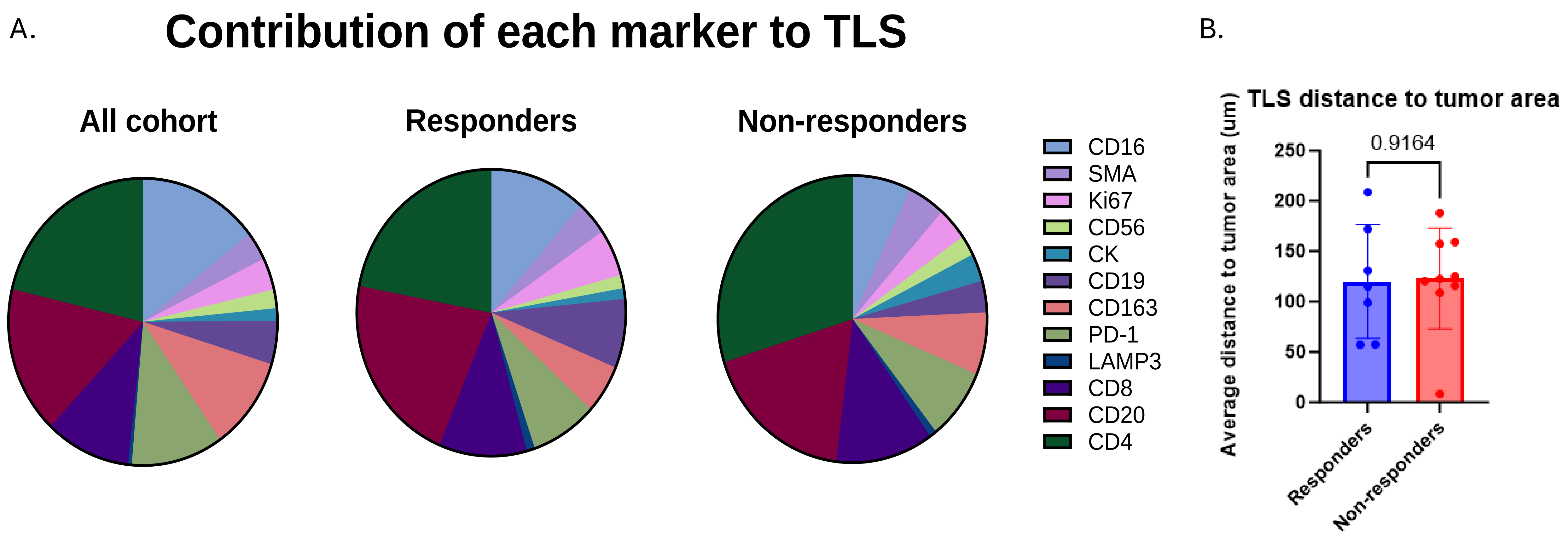


**Supplementary Figure 5.** Survival outcomes based on average TLS size

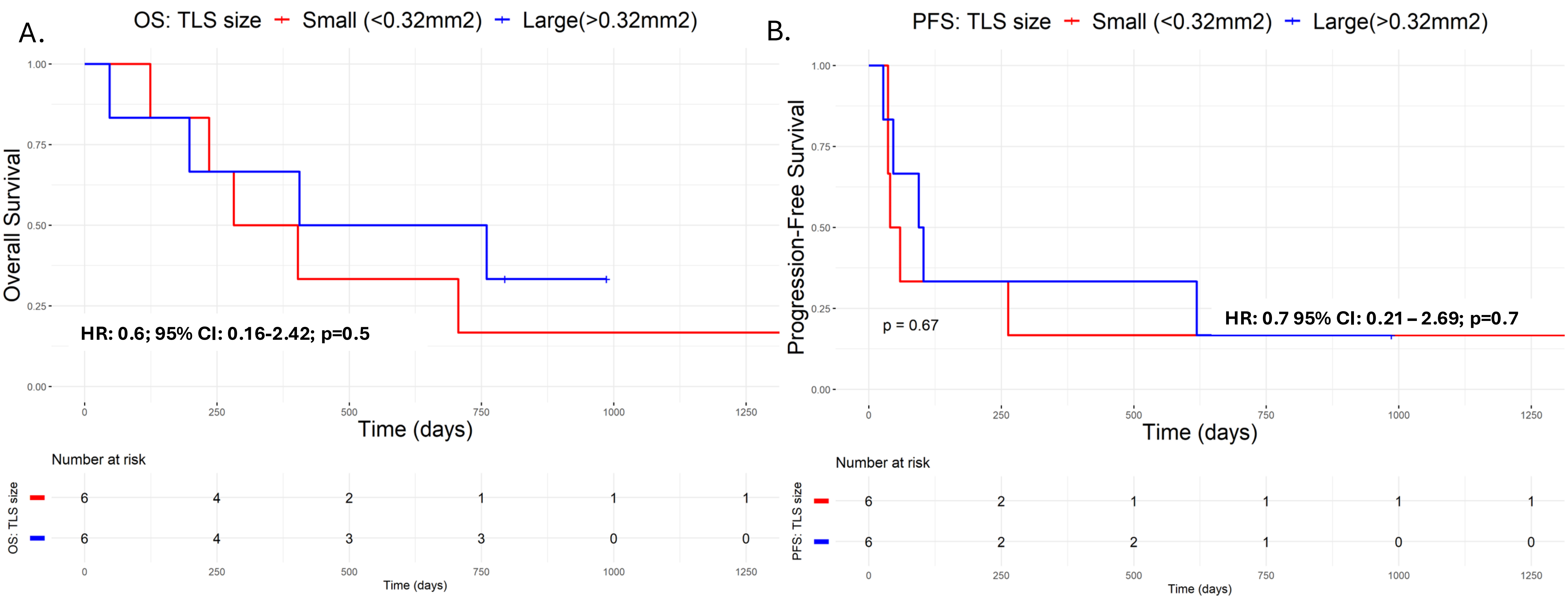
