## Supplementary Figure 6 and 7 for "Immune Cell Densities Predict Response to Immune Checkpoint-Blockade in Head and Neck Cancer"

**Supplementary Figure 6.** Ratio of densities used to predict Response, Progression Free Survival, and Overall Survival

A.

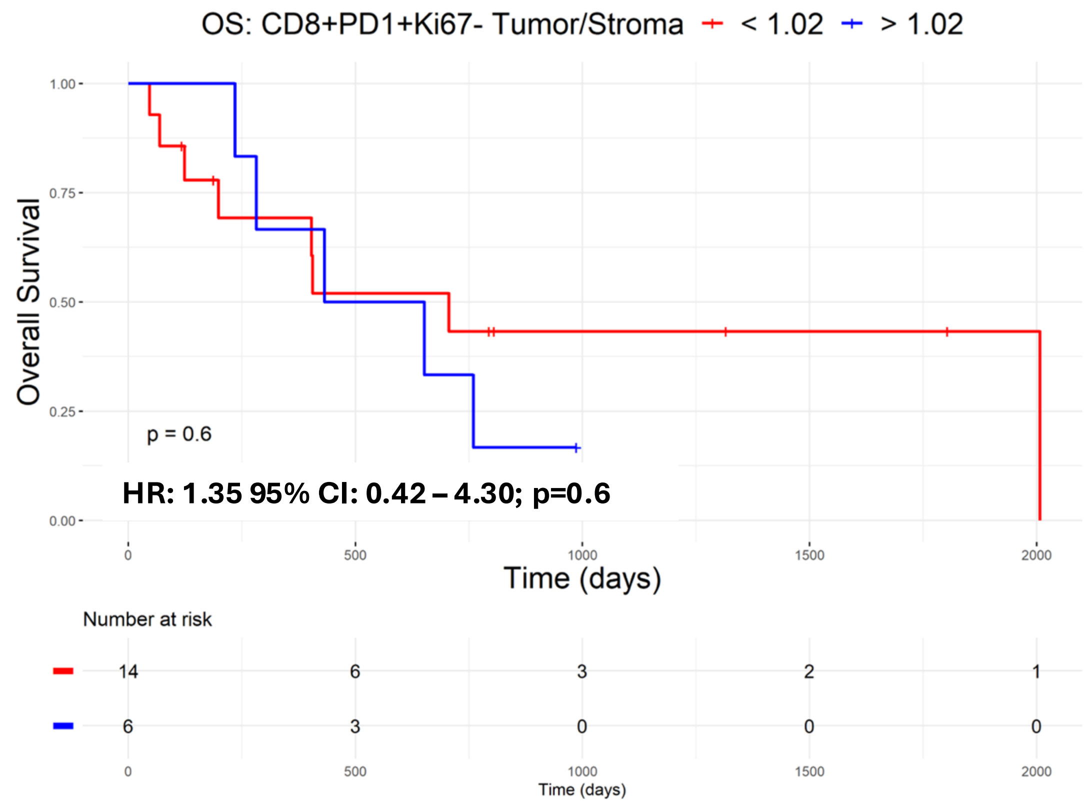


B.

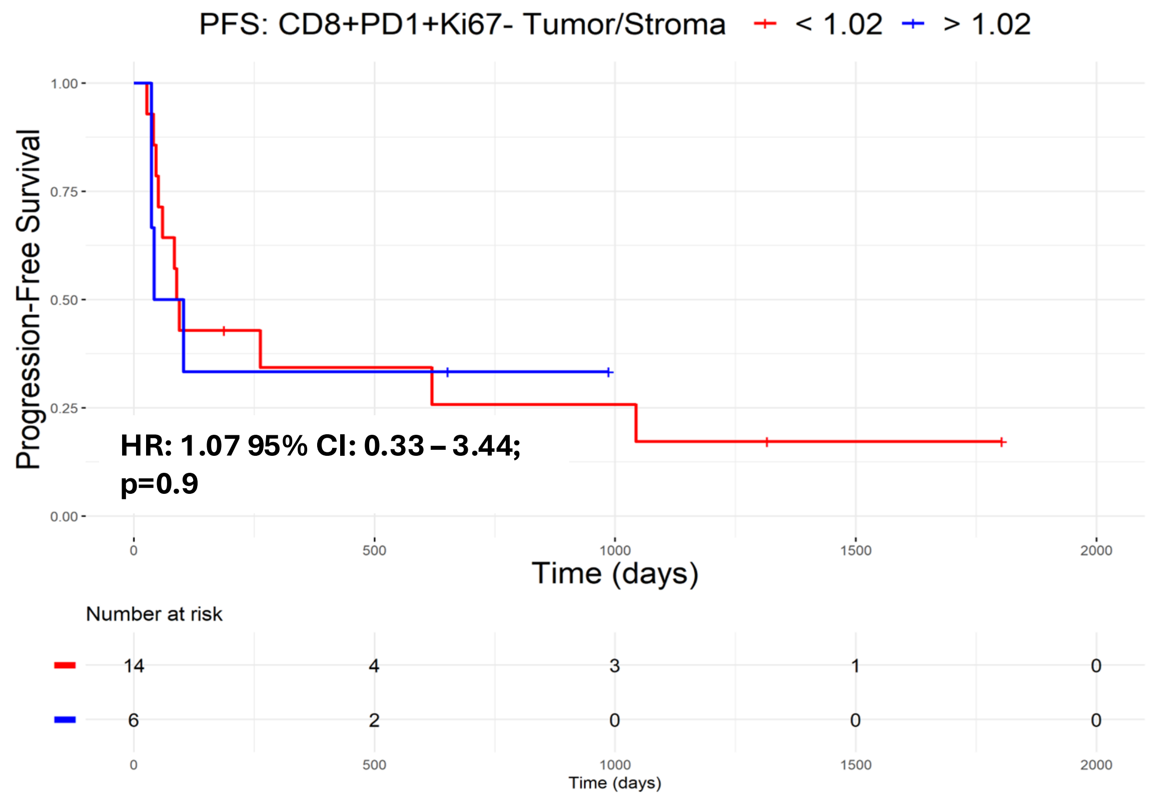


C. Representative mIF image showing strong presence of exhausted T cells in the tumor area

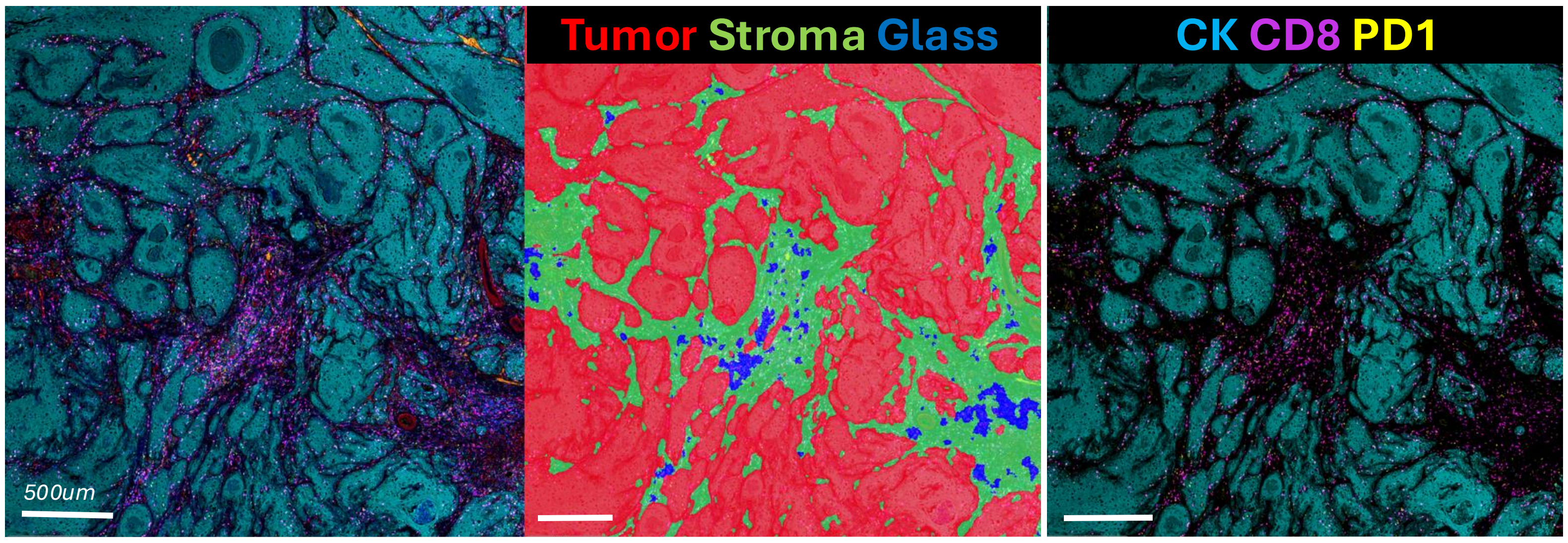


**Supplementary Figure 7.** Ratio of CD20/CD163 densities trend to improve PFS and OS

A.

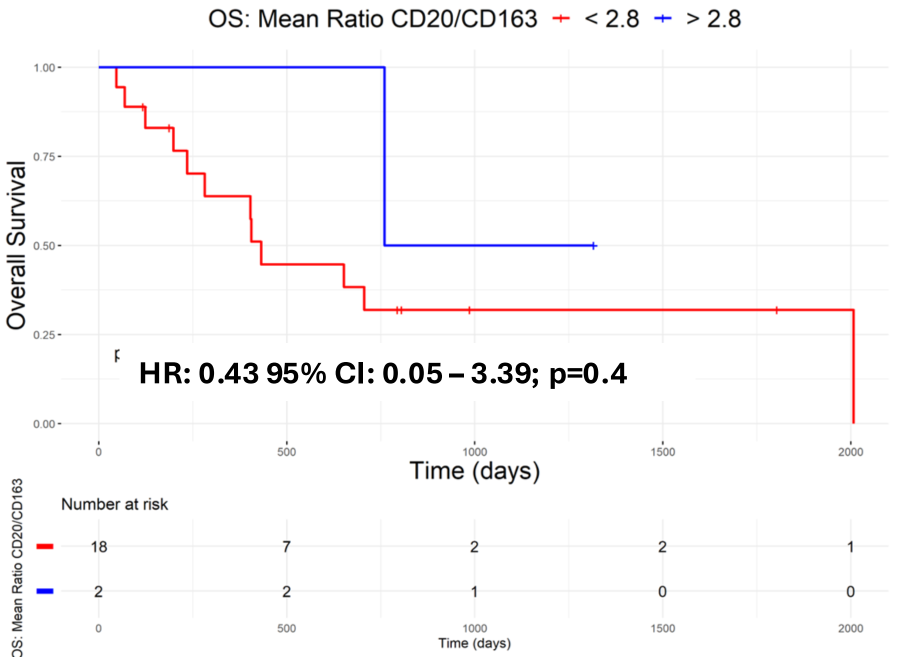


B.

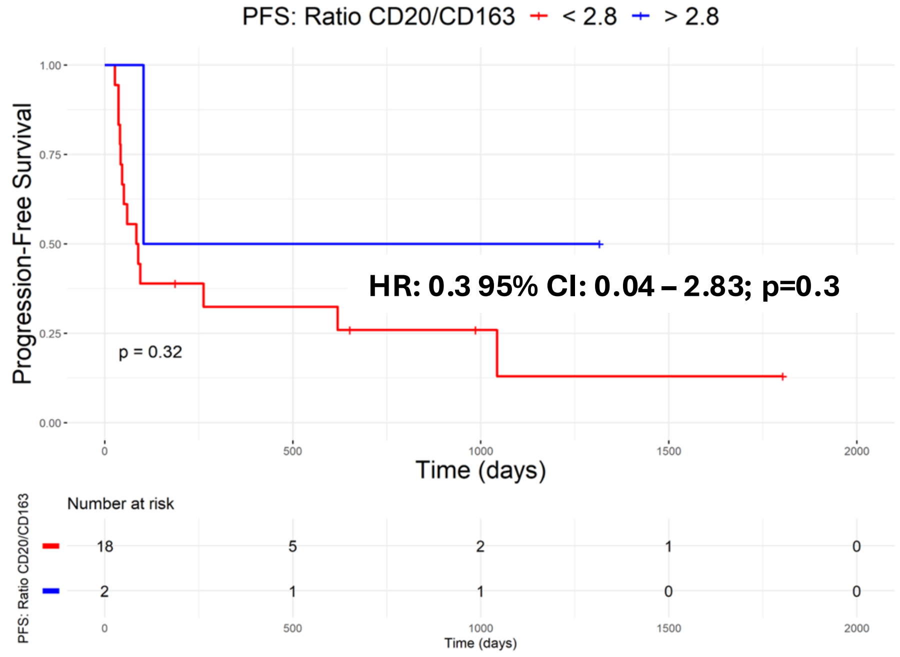


C.

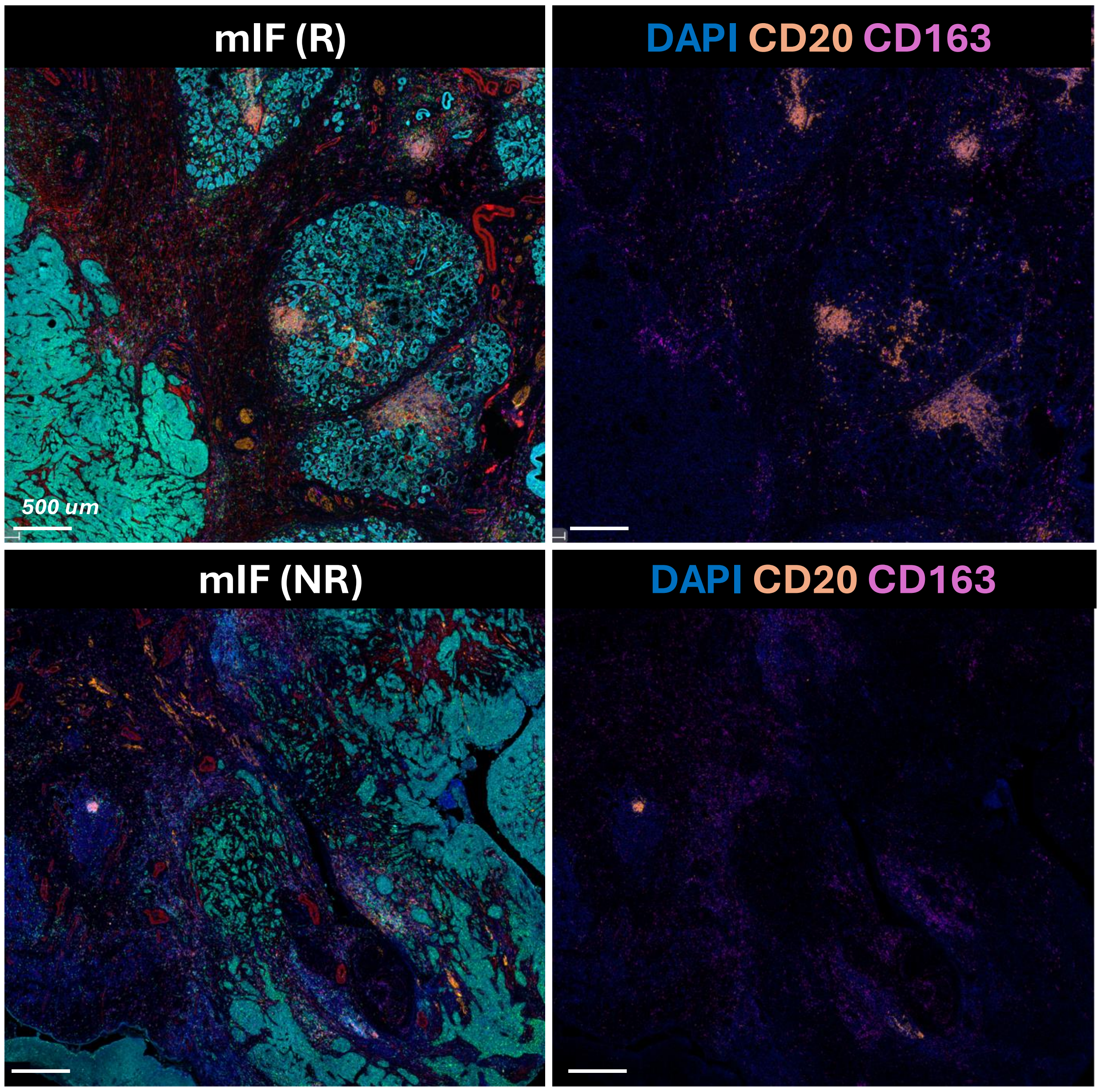
