## Supplementary Figure 8 and 9 for "Immune Cell Densities Predict Response to Immune Checkpoint-Blockade in Head and Neck Cancer"

**Supplementary Figure 8.** Feature Importance for Multiclass Classifiers

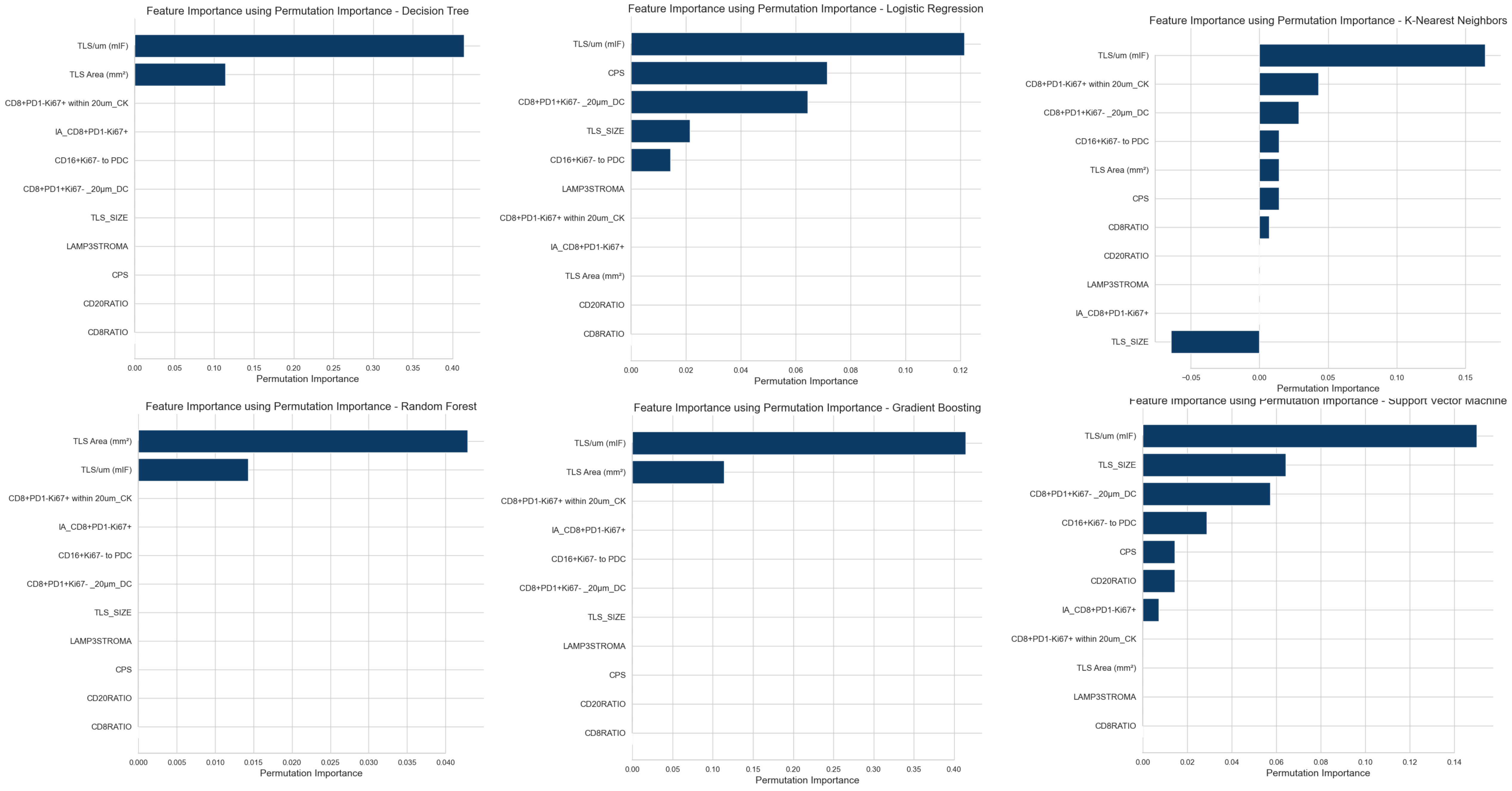


**Supplementary Figure 9.** Presence or absence of TLS and clinical outcomes

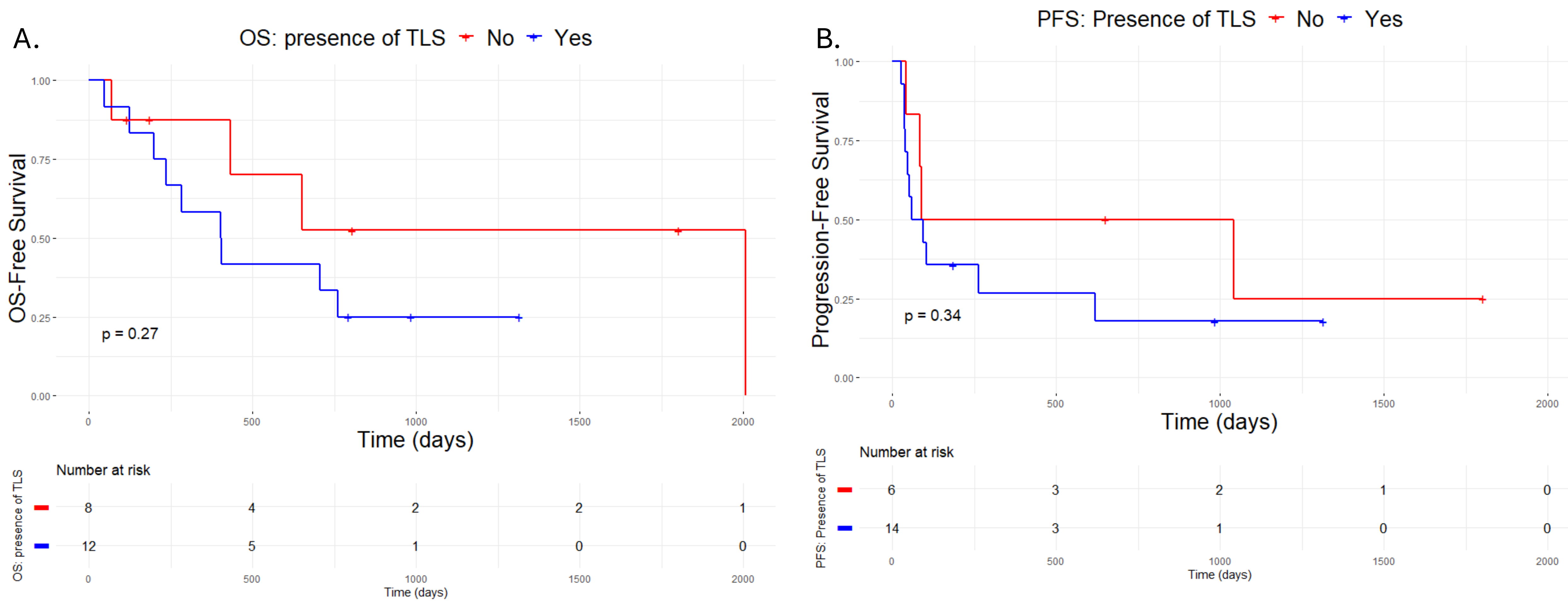
