## Supplementary Table 1 for "Immune Cell Densities Predict Response to Immune Checkpoint-Blockade in Head and Neck Cancer"

**Supplementary Table 1. Staining protocol for panel A, B. A fully custom protocol was optimized using pu**

| | Antibody | Vendor/Clone/Isotype/Lot | Dilution | Incubation Time (min) | Secondary Antibody vendor/lot | Opal/ dilution |
|---|---|---|---|---|---|---|
| Immune Panel (A) | Cytokeratin | Leica/(AE3/AE3)/Mouse-IgG1/6090937 | 1:500 | 30 | Perkin Elmer HRP polymer Opal Ms+ Rb/ARH1001EA | 650/1:100 |
| | CD19 | Leica/BT51E/Mouse-IgG2B/6085347 | 1:50 | 20 | Dako EnVision Secondary. Antibody anti-mouse/K4001/LOT: 10136195 | CF608R/1:50 |
| | CD56 | Leica/CD564/Mouse/6081827 | 1:100 | 20 | Perkin Elmer HRP polymer Opal Ms+ Rb/ARH1001EA | 620/ 1:100 |
| | Ki67 | Thermo Fisher/SP6/Rabbit-IgG/WF3303941 | 1:300 | 20 | Perkin Elmer HRP polymer Opal Ms+ Rb/ARH1001EA | 570/ 1:250 |
| | Alpha-SMA | Dako/1A4/Mouse-IgG2a, kappa/41547637 | 1:100 | 20 | Perkin Elmer HRP polymer Opal Ms+ Rb/ARH1001EA | 540/ 1:250 |

| | | | | | | |
|---|---|---|---|---|---|---|
| | CD16 | Cell Signaling/D1N9L/Rabbit IgG/24326 | 1:200 | 20 | Perkin Elmer HRP polymer Opal Ms+ Rb/ARH1001EA | 520/ 1:100 |
| | DAPI | Spectral DAPI FP1490 | 15 ul/mL | 2 | | - |

| | Antibody | Vendor/Clone/Isotype | Dilution | Incubation Time (min) | Secondary Antibody vendor/lot | Opal/ dilution |
|---|---|---|---|---|---|---|
| | PD1 | Abcam/EPR4877/ Rabbit monoclonal-IgG/GR3230470-4 | 1:500 | 30 | Perkin Elmer HRP polymer Opal Ms+ Rb/ARH1001EA | 520/1:100 |
| | CD163 | Leica/10D6/Mouse/6090602 | 1:500 | 20 | Dako EnVision Secondary. Antibody anti-mouse/K4001/LOT: 10136195 | CF680R/1:50 |
| TLS panel (B) | CD4 | Abcam/EPR6855/ Rabbit monoclonal-IgG/GR3276764-14 | 1:500 | 20 | Perkin Elmer HRP polymer Opal Ms+ Rb/ARH1001EA | 650/ 1:250 |

| | | | | | | |
|---|---|---|---|---|---|---|
| | LAMP3 | Thermo Fisher/ (PA5-84069) Polyclonal-Rabbit-IgG/XA3488279B | 1:300 | 30 | Perkin Elmer HRP polymer Opal Ms+ Rb/ARH1001EA | 540/ 1:150 |
| | CD8 | Leica/4B11/ Mouse-IgG2b/6070827 | 1:300 | 20 | Perkin Elmer HRP polymer Opal Ms+ Rb/ARH1001EA | 570/ 1:250 |
| | CD20 | Leica/L26/Mouse-IgG2A, kappa/6070984 | 1:500 | 20 | Perkin Elmer HRP polymer Opal Ms+ Rb/ARH1001EA | 620/ 1:100 |
| | DAPI | Spectral DAPI FP1490 | 15 ul/mL | 2 | | - |

**ıblicly available mIF staining guidelines (Akoya). Low-expressed targets were paired with more potent Op**

| Inuobation time (min) | Position in the reaction |
|---|---|
| 10 | 1 |
| 10 | 2 |
| 10 | 3 |
| 10 | 4 |
| 10 | 5 |

| | |
|---|---|
| 10 | 6 |
| | 7 |

| | Position in the reaction |
|---|---|
| 10 | 1 |
| 10 | 2 |
| 10 | 3 |

| | |
|---|---|
| 10 | 4 |
| 10 | 5 |
| 10 | 6 |
| | 7 |

**[illegible]als. Of note, CF680R was used instead of Opal 690 due to improved spectral behavior. Antigen Retrieval [illegible]**

**Citrate pH 6 buffer was used for all antibodies but for CD20, in which ARB pH 9 EDTA was used.**
