## Supplementary Table 2 for "Immune Cell Densities Predict Response to Immune Checkpoint-Blockade in Head and Neck Cancer"

**Supplementary Table 2. HALO phenotyping parameters. Parameters for phenotyping of cells in spatial analysis software (Halo Indica Labs). The same parameters were applied to every sample to phenotype each cell.**

| Variable | Value |
|---|---|
| **Nuclear detection** | |
| Nuclear contrast threshod | 0.499 |
| Minimum Nuclear Intensity | 0.04 |
| Maximum Image Brightness | 0.974 |
| Nuclear Segmentation Aggressiveness | 0.841 |
| Fill Nuclear Holes | FALSE |
| Nuclear size | 10,539.82 |
| Minimum nuclear Roundness | 0.022 |
| **Membrane and Cytoplasm Detection** | |
| Maximum Cytoplasm Radius | 2.79 |
| Membrane Segmentation Aggressiveness | 0.649 |
| Cell Size | 0,2668.1416 |
| **rker-Opal detection cytoplasm positive threshold*** | |
| Panel A (Imne) | |
| SMA-Opal 540 | 3.9759,69,69 |
| CD19-Opal 680R | 0.7211,11,11 |
| Cytokeratin-Opal 650 | 2.1607,59,59 |
| CD56-Opal 620 | 3.2318,84,84 |
| CD16-Opal 520 | 2.4487,30,30 |
| Ki67-Opal 570 (Nuclear Threshold) | 7.5949,83,83 |
| Panel B (TLS) | |
| PD1-Opal 520 | 0.5037,14,14 |
| LAMP3-Opal 540 | 0.7465,26,26 |
| CD8-Opal 570 | 2.5816,77,77 |
| CD20-Opal 620 | 10.6488,200,200 |
| CD163-Opal 690 | 0.7864,10,10 |
| CD163-Opal 650 | 4,87,87 |
| *Cytoplasm % completeness Threshold was 30% for all channels | |
