## Supplementary Table 3 for "Immune Cell Densities Predict Response to Immune Checkpoint-Blockade in Head and Neck Cancer"

Supplementary Table 3. Spatial analysis

| Average distance to CK+ cells | Responders | Non-responders | p-value* |
|---|---|---|---|
| LAMP3+ | 58.64 (12.4-101.57) | 103.24 (12.46-640.59) | 0.39 |
| CD20+ | 156.34 (13.85-634.87) | 107.97 (277.29-1403.67) | 0.38 |
| SMA+ | 148.09 (20.19-348.13) | 110.85 (20.4-413.96) | 0.42 |
| CD163+ | 127.53 (18.87-226.78) | 130.75 (15.13-696.94) | 0.95 |
| PD1+ | 89.79 (11-238.3) | 103.35 (12.4-535.94) | 0.77 |
| CD8+ | 82.19 (9.22-212.6) | 73.75 (10.66 - 328.91) | 0.72 |
| CD16+ | 63.04 (15.28-135.12) | 72.70 (14.41-267.59) | 0.7 |
| CD4+ | 87.44 (11.09-263.28) | 67.40 (11.91-219.86) | 0.43 |
| CD4+PD1+Ki67- | 87.2 (10.24-246.28) | 63.7 (11.68-184.17) | 0.33 |
| CD4+PD1+Ki67+ | 50.47 (10.24-113.69) | 47.4 (6.94-188.03) | 0.86 |
| CD4+PD1-Ki67+ | 52.55 (10.26-128.56) | 37.94 (9.95-131.72) | 0.3 |
| CD8+PD1+Ki67- | 85.5 (8.75-296.08) | 54.2 (10.8-195.77) | 0.3 |
| CD8+PD1+Ki67+ | 36.98 (7.58-106.89) | 34.44 (7.99-155.19) | 0.86 |
| CD8+PD1-Ki67+ | 54.5 (8.25-118.7) | 48.68 (8.62-233.03) | 0.78 |
| Average distance of LAMP3+Ki67-PD1- to: | Responders | Non-responders | p-value |
| CD4+PD1+Ki67- | 198.18 (48.77-752.32) | 238.89 (32.06-687.05) | 0.66 |
| CD4+PD1+Ki67+ | 798.73 (150.04-5028.47) | 942.08 (107.36-3031.19) | 0.69 |

| | | | |
|---|---|---|---|
| CD4+PD1-Ki67+ | 237.20 (68.72-859.56) | 211.56 (66.24-416.55) | 0.76 |
| CD8+PD1+Ki67 | 213.67 (45-772.63) | 277.82 (38.1-1417.65) | 0.62 |
| CD8+PD1+Ki67 | 838.32 (127.56-43550.05) | 679.80 (154.31-1926.18) | 0.7 |
| CD8+PD1-Ki67 | 323.06 (76.05-1264.67) | 278.71 (76.33-669.21) | 0.72 |
| Average distance of x marker to LAMP3+Ki67+ | Responders | Non-responders | p-value |
| CD16+Ki67- to PDC | 635.69 (180.55-1920.96) | 1327.56 (166.86-5010.69) | 0.14 |
| CD16+Ki67+ to PDC | 596.99 (154.75-1864.52) | 1255.86 (138.29-4572.34) | 0.16 |
| CD4+PD1+Ki67- to PDC | 773.36 (237.36-2179.86) | 1268.55 (122.07-5690.4) | 0.34 |
| CD4+PD1+Ki67+ to PDC | 681.57 (151.19-2010.21) | 1087.46 (81.36-3683.8) | 0.3 |
| CD4+PD1-Ki67+ to PDC | 715.39 (158.42-2124.98) | 1272.69 (132.73-5000.11) | 0.26 |
| CD8+PD1+Ki67- to PDC | 826.33 (218.17-2251.46) | 1259.85 (124.64-5468.3) | 0.37 |
| CD8+PD1+Ki67+ to PDC | 926.71 (131.75-4259.8) | 1086.68 (80.67-4091.45) | 0.74 |
| CD8+PD1-Ki67 | 734.33 (167.75-2056.17) | 1179.27 (113.99-4722.03) | 0.33 |
| Average distance to tumor/stroma interface | Responders | Non-responders | p-value |
| CD8 | 41.44 (-164.41; 175.45) | 47.13 (-109.71; 199.35) | 0.86 |
| CD4 | 58.55 (-125.54; 141.06) | 60.52 (-12.61; 138.75) | 0.93 |
| PD1 | 41.47 (-206.48; 129.91) | 44.02 (-26.02; 139.26) | 0.93 |
| CD20 | 133.22 (7.86; 243.53) | 130.58 (19.46; 228.56) | 0.92 |

| | | | |
|---|---|---|---|
| CD16 | 11.12 (-181.36; 102.93) | 52.31 (-28.83; 194.09) | 0.22 |
| CD163 | 97.61 (-89.16; 215.85) | 96.411 (-21.56; 243.15) | 0.97 |
| LAMP3 | -3.74 ( -173.82; 145.87) | 60.30 (-28.25 - 144.38) | 0.059 |
| SMA | 111.53 (-84.04; 199.47) | 104.57 (24.06-199.95) | 0.79 |
| CD4+PD1+Ki67- | 112.88 (20.56-175.72) | 98.05 (38.09-152.79) | 0.43 |
| CD4+PD1+Ki67+ | 106.4 (23.54-252.44) | 80.109 (9.68-156.53) | 0.24 |
| CD4+PD1-Ki67+ | 86.06 (22.11-172.49) | 71.85 (19.65-162.46) | 0.41 |
| CD8+PD1+Ki67- | 103.98 (19.87-191.17) | 85.54 (29.48-138.3) | 0.25 |
| CD8+PD1+Ki67+ | 74.35 (0-149.64) | 55.83 (9.12-117.28) | 0.29 |
| CD8+PD1-Ki67+ | 88.67 (17.37-160.44) | 71.07 (15.72-178.77) | 0.33 |
| CD16+Ki67+ | 82.95 (28.05-133.14) | 72.49 (27.15-199.38) | 0.56 |
| CD16+KI67- | 98.64 (21.01-172.08) | 87.79 (35.08-207.56) | 0.56 |
| LAMP3+Ki67+ | 90.36 (20.14-151.( | 76.33 (0-187.15) | 0.46 |
| *unpaired t-test | | | |
