## Supplementary Table 4 for "Immune Cell Densities Predict Response to Immune Checkpoint-Blockade in Head and Neck Cancer"

**Supplementary Table 4. Density analysis**

| **CD4+PD1+Ki67-_tumor** | | t-Test: Two-Sample Assuming Equal Variar | | |
|---|---|---|---|---|
| R | NR | | | |
| 0.000485 | 0.012641 | | *Variable 1* | *Variable 2* |
| 0.00607 | 0.044458 | Mean | 0.05357091 | 0.02509115 |
| 0.023859 | 0.01731 | Variance | 0.00390976 | 0.00090665 |
| 0.063136 | 9.60E-05 | Observations | 11 | 13 |
| 0.221281 | 0.008648 | Pooled Varian | 0.0022717 | |
| 0.024517 | 0.000589 | Hypothesized | 0 | |
| 0.064851 | 0.003685 | df | 22 | |
| 0.078862 | 0.028266 | t Stat | 1.4585557 | |
| 0.028858 | 0.000136 | P(T<=t) one-ta | 0.07940878 | |
| 0.005087 | 0.100121 | t Critical one- | 1.71714437 | |
| 0.072274 | 0.033785 | P(T<=t) two-ta | 0.15881755 | |
| | 0.00948 | t Critical two-t | 2.07387307 | |
| | 0.06697 | | | |

| **CD4+PD1-Ki67-_Stroma** | | t-Test: Two-Sample Assuming Equal Variar | | |
|---|---|---|---|---|
| R | NR | | | |
| 0.204982 | 0.142852 | | *Variable 1* | *Variable 2* |
| 0.148165 | 0.577378 | Mean | 0.56917991 | 0.333434 |
| 0.50134 | 0.1493 | Variance | 0.15617516 | 0.0635741 |
| 0.497048 | 0.231334 | Observations | 11 | 13 |
| 1.22134 | 0.166188 | Pooled Varian | 0.10566549 | |
| 0.599155 | 0.066877 | Hypothesized | 0 | |
| 0.972232 | 0.146382 | df | 22 | |
| 1.19241 | 0.354751 | t Stat | 1.77027188 | |
| 0.317415 | 0.132267 | P(T<=t) one-ta | 0.04527007 | |
| 0.47717 | 0.784335 | t Critical one- | 1.71714437 | |
| 0.129722 | 0.449067 | P(T<=t) two-ta | 0.09054015 | |
| | 0.813985 | t Critical two-t | 2.07387307 | |
| | 0.319926 | | | |

| **CD4+PD1-Ki67-_Tumor** | | t-Test: Two-Sample Assuming Equal Variar | | |
|---|---|---|---|---|
| R | NR | | | |
| 0.018446 | 0.050825 | | *Variable 1* | *Variable 2* |
| 0.034325 | 0.087044 | Mean | 0.17963245 | 0.10516654 |
| 0.063312 | 0.051941 | Variance | 0.01860455 | 0.01013266 |
| 0.232184 | 0.017205 | Observations | 11 | 13 |
| 0.456476 | 0.131532 | Pooled Varian | 0.01398351 | |
| 0.154057 | 0.017323 | Hypothesized | 0 | |
| 0.257368 | 0.05736 | df | 22 | |
| 0.243668 | 0.105711 | t Stat | 1.53713411 | |

| | | | | |
|---|---|---|---|---|
| 0.101258 | 0.010954 | P(T<=t) one-ta | 0.06925972 | |
| 0.090107 | 0.336877 | t Critical one- | 1.71714437 | |
| 0.324756 | 0.285021 | P(T<=t) two-ta | 0.13851945 | |
| | 0.140219 | t Critical two-t | 2.07387307 | |
| | 0.075153 | | | |
| **CD8+PD1+Ki67-_Stroma** | | t-Test: Two-Sample Assuming Equal Variar | | |
| R | NR | | | |
| 0.005099 | 0.013149 | | *Variable 1* | *Variable 2* |
| 0.019272 | 0.234596 | Mean | 0.12408282 | 0.05216215 |
| 0.126825 | 0.015704 | Variance | 0.05225555 | 0.00570656 |
| 0.111664 | 0.001433 | Observations | 11 | 13 |
| 0.801374 | 0.005984 | Pooled Varian | 0.02686519 | |
| 0.084362 | 0.005567 | Hypothesized | 0 | |
| 0.06429 | 0.031328 | df | 22 | |
| 0.059587 | 0.027928 | t Stat | 1.07107809 | |
| 0.076365 | 0.000887 | P(T<=t) one-ta | 0.14787227 | |
| 0.009585 | 0.126463 | t Critical one- | 1.71714437 | |
| 0.006488 | 0.023365 | P(T<=t) two-ta | 0.29574454 | |
| | 0.019529 | t Critical two-t | 2.07387307 | |
| | 0.172175 | | | |
| **CD8+PD1+Ki67-_Tumor** | | t-Test: Two-Sample Assuming Equal Variar | | |
| R | NR | | | |
| 0.000757 | 0.053576 | | *Variable 1* | *Variable 2* |
| 0.002018 | 0.123442 | Mean | 0.06266364 | 0.03688069 |
| 0.047604 | 0.030494 | Variance | 0.00580993 | 0.00135238 |
| 0.124575 | 0.000331 | Observations | 11 | 13 |
| 0.263007 | 0.013162 | Pooled Varian | 0.00337854 | |
| 0.048964 | 0.004692 | Hypothesized | 0 | |
| 0.025806 | 0.003263 | df | 22 | |
| 0.036842 | 0.046532 | t Stat | 1.08275565 | |
| 0.090458 | 0.000432 | P(T<=t) one-ta | 0.14532103 | |
| 0.008606 | 0.035314 | t Critical one- | 1.71714437 | |
| 0.040663 | 0.037229 | P(T<=t) two-ta | 0.29064206 | |
| | 0.040093 | t Critical two-t | 2.07387307 | |
| | 0.090889 | | | |
| **CD8+PD1-Ki67-_Stroma** | | t-Test: Two-Sample Assuming Equal Variar | | |
| R | NR | | | |
| 0.081914 | 0.064994 | | *Variable 1* | *Variable 2* |
| 0.048727 | 0.220625 | Mean | 0.35341927 | 0.17633138 |
| 0.280649 | 0.054826 | Variance | 0.14393642 | 0.02230368 |
| 0.18662 | 0.095487 | Observations | 11 | 13 |

| | | | | |
|---|---|---|---|---|
| 1.2689 | 0.067509 | Pooled Varian | 0.07759129 | |
| 0.13705 | 0.026713 | Hypothesized | 0 | |
| 0.188786 | 0.282834 | df | 22 | |
| 0.33411 | 0.226289 | t Stat | 1.55183223 | |
| 0.561436 | 0.070582 | P(T<=t) one-ta | 0.0674851 | |
| 0.773702 | 0.107088 | t Critical one- | 1.71714437 | |
| 0.025718 | 0.293998 | P(T<=t) two-ta | 0.13497021 | |
| | 0.567041 | t Critical two-t | 2.07387307 | |
| | 0.214322 | | | |
| **CD8+PD1-Ki67-_Tumor** | | t-Test: Two-Sample Assuming Equal Variar | | |
| R | NR | | | |
| 3.00E-05 | 0.008982 | | *Variable 1* | *Variable 2* |
| 0.000224 | 0.020881 | Mean | 0.01887473 | 0.02401823 |
| 0.009714 | 0.003746 | Variance | 0.00047203 | 0.00380377 |
| 0.02135 | 6.90E-05 | Observations | 11 | 13 |
| 0.075111 | 0.006921 | Pooled Varian | 0.00228935 | |
| 0.006609 | 0.000914 | Hypothesized | 0 | |
| 0.020093 | 0.000758 | df | 22 | |
| 0.015653 | 0.013438 | t Stat | -0.2624011 | |
| 0.036154 | 0.000125 | P(T<=t) one-ta | 0.39772685 | |
| 0.001584 | 0.008474 | t Critical one- | 1.71714437 | |
| 0.0211 | 0.00482 | P(T<=t) two-ta | 0.7954537 | |
| | 0.014929 | t Critical two-t | 2.07387307 | |
| | 0.22818 | | | |
| **LAMP3+PD1+Ki67-_Stroma** | | t-Test: Two-Sample Assuming Equal Variar | | |
| R | NR | | | |
| 9.50E-05 | 0.014122 | | *Variable 1* | *Variable 2* |
| 0.000722 | 0.003324 | Mean | 0.02318964 | 0.01568354 |
| 0.04411 | 0.026682 | Variance | 0.00077432 | 0.0008549 |
| 0.087658 | 0.004372 | Observations | 11 | 13 |
| 0.018157 | 0.000218 | Pooled Varian | 0.00081827 | |
| 0.001492 | 0.000345 | Hypothesized | 0 | |
| 0.053061 | 0.023334 | df | 22 | |
| 0.023295 | 0.006183 | t Stat | 0.64051331 | |
| 0.01715 | 1.10E-05 | P(T<=t) one-ta | 0.26422732 | |
| 0.005213 | 0.003644 | t Critical one- | 1.71714437 | |
| 0.004133 | 0.012624 | P(T<=t) two-ta | 0.52845464 | |
| | 0.000588 | t Critical two-t | 2.07387307 | |
| | 0.108439 | | | |
| **LAMP3+PD1+Ki67-_Stroma** | | t-Test: Two-Sample Assuming Equal Variar | | |
| R | NR | | | |

| | | | | |
|---|---|---|---|---|
| 0.000212 | 0.002712 | | *Variable 1* | *Variable 2* |
| 0.000238 | 0.000225 | Mean | 0.00497855 | 0.007703 |
| 0.008436 | 0.017021 | Variance | 2.1989E-05 | 0.0002325 |
| 0.014604 | 0.003596 | Observations | 11 | 13 |
| 0.004237 | 0.000163 | Pooled Varian | 0.00013681 | |
| 0.000138 | 0.000183 | Hypothesized | 0 | |
| 0.008515 | 0.007014 | df | 22 | |
| 0.00786 | 0.002752 | t Stat | -0.5685645 | |
| 0.003505 | 0 | P(T<=t) one-ta | 0.28770513 | |
| 0.000442 | 0.000869 | t Critical one- | 1.71714437 | |
| 0.006577 | 0.003935 | P(T<=t) two-ta | 0.57541025 | |
| | 0.005569 | t Critical two-t | 2.07387307 | |
| | 0.0561 | | | |
| **LAMP3+PD1-Ki67-_Stroma** | | t-Test: Two-Sample Assuming Equal Variar | | |
| R | NR | | | |
| 0.001975 | 0.010199 | | *Variable 1* | *Variable 2* |
| 0.001156 | 0.002605 | Mean | 0.01587373 | 0.01309823 |
| 0.006557 | 0.01185 | Variance | 0.00027789 | 0.00022398 |
| 0.015507 | 0.006715 | Observations | 11 | 13 |
| 0.009893 | 0.008418 | Pooled Varian | 0.00024848 | |
| 0.002307 | 0.000406 | Hypothesized | 0 | |
| 0.036157 | 0.020644 | df | 22 | |
| 0.052848 | 0.008932 | t Stat | 0.4297878 | |
| 0.029817 | 0.001183 | P(T<=t) one-ta | 0.33576516 | |
| 0.009533 | 0.003262 | t Critical one- | 1.71714437 | |
| 0.008861 | 0.044356 | P(T<=t) two-ta | 0.67153033 | |
| | 0.006768 | t Critical two-t | 2.07387307 | |
| | 0.044939 | | | |
| **LAMP3+PD1-Ki67-_Tumor** | | t-Test: Two-Sample Assuming Equal Variar | | |
| R | NR | | | |
| 0.04008 | 0.007929 | | *Variable 1* | *Variable 2* |
| 0.001711 | 0.000156 | Mean | 0.02133409 | 0.00755408 |
| 0.002062 | 0.002302 | Variance | 0.00095251 | 9.4599E-05 |
| 0.003483 | 0.000331 | Observations | 11 | 13 |
| 0.002345 | 0.007125 | Pooled Varian | 0.00048456 | |
| 0.001079 | 0.000122 | Hypothesized | 0 | |
| 0.039718 | 0.004278 | df | 22 | |
| 0.102247 | 0.007189 | t Stat | 1.52805451 | |
| 0.01238 | 0.000133 | P(T<=t) one-ta | 0.07037506 | |
| 0.003019 | 0.001272 | t Critical one- | 1.71714437 | |
| 0.026551 | 0.030891 | P(T<=t) two-ta | 0.14075011 | |

| | | | | |
|---|---|---|---|---|
| | 0.024019 | t Critical two-t | 2.07387307 | |
| | 0.012456 | | | |
| **LAMP3+PD1-Ki67+_Stroma** | | t-Test: Two-Sample Assuming Equal Variar | | |
| R | NR | | | |
| 0.000151 | 0.000171 | | *Variable 1* | *Variable 2* |
| 2.40E-05 | 0.000121 | Mean | 0.00145409 | 0.00070885 |
| 0.000298 | 0.000709 | Variance | 3.488E-06 | 1.0622E-06 |
| 0.000748 | 0.000317 | Observations | 11 | 13 |
| 0.000695 | 0.000508 | Pooled Varian | 2.1648E-06 | |
| 8.80E-05 | 0 | Hypothesized | 0 | |
| 0.002132 | 0.000798 | df | 22 | |
| 0.005715 | 0.00049 | t Stat | 1.23637161 | |
| 0.004215 | 2.30E-05 | P(T<=t) one-ta | 0.11467905 | |
| 0.000698 | 0.000381 | t Critical one-t | 1.71714437 | |
| 0.001231 | 0.001335 | P(T<=t) two-ta | 0.22935809 | |
| | 0.000441 | t Critical two-t | 2.07387307 | |
| | 0.003921 | | | |
| **LAMP3+PD1-Ki67+_Tumor** | | t-Test: Two-Sample Assuming Equal Variar | | |
| R | NR | | | |
| 0.000636 | 0.000761 | | *Variable 1* | *Variable 2* |
| 5.90E-05 | 9.00E-06 | Mean | 0.00928345 | 0.00279877 |
| 0.000597 | 0.000651 | Variance | 0.00027447 | 1.8925E-05 |
| 0.000697 | 1.40E-05 | Observations | 11 | 13 |
| 0.000453 | 0.001305 | Pooled Varian | 0.00013508 | |
| 0.000113 | 0 | Hypothesized | 0 | |
| 0.044565 | 0.000426 | df | 22 | |
| 0.039887 | 0.006013 | t Stat | 1.36192637 | |
| 0.005018 | 0 | P(T<=t) one-ta | 0.09350251 | |
| 0.000682 | 0.000254 | t Critical one-t | 1.71714437 | |
| 0.009411 | 0.003558 | P(T<=t) two-ta | 0.18700503 | |
| | 0.010364 | t Critical two-t | 2.07387307 | |
| | 0.013029 | | | |
| **CD20** | | t-Test: Two-Sample Assuming Equal Variar | | |
| R | NR | | | |
| 0.036296 | 0.068522 | | *Variable 1* | *Variable 2* |
| 0.322226 | 0.527542 | Mean | 0.59237655 | 0.15777769 |
| 0.766277 | 0.012346 | Variance | 0.37025921 | 0.04864192 |
| 0.835833 | 0.173046 | Observations | 11 | 13 |
| 0.077475 | 0.057798 | Pooled Varian | 0.1948316 | |
| 1.09582 | 0.036296 | Hypothesized | 0 | |
| 0.739297 | 0.087734 | df | 22 | |

| | | | | |
|---|---|---|---|---|
| 0.013696 | 0.412863 | t Stat | 2.40337357 | |
| 2.00878 | 0.000528 | P(T<=t) one-ta | 0.01255659 | |
| 0.001664 | 0.000884 | t Critical one- | 1.71714437 | |
| 0.618778 | 0.640747 | P(T<=t) two-ta | **0.02511318** | |
| | 0.029096 | t Critical two-t | 2.07387307 | |
| | 0.003708 | | | |
| **PD1** | | t-Test: Two-Sample Assuming Equal Variar | | |
| R | NR | | | |
| 0.371999 | 0.156323 | | *Variable 1* | *Variable 2* |
| 0.737748 | 0.125566 | Mean | 0.74351 | 0.69820885 |
| 0.476727 | 0.070218 | Variance | 1.01100488 | 1.16414929 |
| 0.678767 | 0.291122 | Observations | 11 | 13 |
| 0.02432 | 0.327876 | Pooled Varian | 1.0945382 | |
| 0.910627 | 0.371999 | Hypothesized | 0 | |
| 0.474463 | 0.424996 | df | 22 | |
| 0.523709 | 2.03837 | t Stat | 0.10569543 | |
| 3.66463 | 0.06109 | P(T<=t) one-ta | 0.45839091 | |
| 0.23442 | 3.66507 | t Critical one- | 1.71714437 | |
| 0.0812 | 1.46778 | P(T<=t) two-ta | 0.91678182 | |
| | 0.070777 | t Critical two-t | 2.07387307 | |
| | 0.005528 | | | |
| **LAMP3** | | | | |
| R | NR | t-Test: Two-Sample Assuming Equal Variar | | |
| 0.06496 | 0.097495 | | | |
| 0.128811 | 0.050032 | | *Variable 1* | *Variable 2* |
| 0.236951 | 0.017773 | Mean | 0.079693 | 0.05621323 |
| 0.19183 | 0.032783 | Variance | 0.00580103 | 0.00769697 |
| 0.04318 | 0.059574 | Observations | 11 | 13 |
| 0.067565 | 0.06496 | Pooled Varian | 0.00683518 | |
| 0.076273 | 0.039727 | Hypothesized | 0 | |
| 0.005317 | 0.010169 | df | 22 | |
| 0.037668 | 0.001077 | t Stat | 0.69323642 | |
| 0.003957 | 0.332044 | P(T<=t) one-ta | 0.24770775 | |
| 0.020111 | 0.006682 | t Critical one- | 1.71714437 | |
| | 0.017094 | P(T<=t) two-ta | 0.49541549 | |
| | 0.001362 | t Critical two-t | 2.07387307 | |
| **CD4** | | | | |
| R | NR | t-Test: Two-Sample Assuming Equal Variar | | |
| 1.02607 | 0.864444 | | | |
| 2.09199 | 1.12989 | | *Variable 1* | *Variable 2* |
| 1.85637 | 0.367917 | Mean | 1.10261345 | 0.63951954 |

| | | | | |
|---|---|---|---|---|
| 0.236707 | 0.595277 | Variance | 0.65946471 | 0.3010218 |
| 0.843949 | 0.257001 | Observations | 11 | 13 |
| 0.616296 | 0.265471 | Pooled Varian | 0.4639504 | |
| 0.769668 | 0.237758 | Hypothesized | 0 | |
| 0.969911 | 1.95694 | df | 22 | |
| 2.81289 | 0.091527 | t Stat | 1.65957047 | |
| 0.240753 | 0.968476 | P(T<=t) one-ta | 0.05559503 | |
| 0.664144 | 1.16344 | t Critical one- | 1.71714437 | |
| | 0.264759 | P(T<=t) two-ta | 0.11119006 | |
| | 0.150854 | t Critical two-t | 2.07387307 | |
| **CD8** | | | | |
| R | NR | t-Test: Two-Sample Assuming Equal Variar | | |
| 0.654809 | 0.737224 | | | |
| 0.625325 | 1.2009 | | *Variable 1* | *Variable 2* |
| 0.368412 | 0.1422 | Mean | 0.86145827 | 0.42152854 |
| 0.098795 | 0.485504 | Variance | 0.86643422 | 0.12740095 |
| 0.565577 | 0.361032 | Observations | 11 | 13 |
| 1.58486 | 0.133395 | Pooled Varian | 0.46332516 | |
| 0.278649 | 0.283096 | Hypothesized | 0 | |
| 0.326439 | 0.341895 | df | 22 | |
| 3.09792 | 0.049574 | t Stat | 1.57762133 | |
| 0.074205 | 0.873958 | P(T<=t) one-ta | 0.06446225 | |
| 1.80105 | 0.684082 | t Critical one- | 1.71714437 | |
| | 0.104678 | P(T<=t) two-ta | 0.12892451 | |
| | 0.082333 | t Critical two-t | 2.07387307 | |
| **CD163** | | | | |
| R | NR | t-Test: Two-Sample Assuming Equal Variar | | |
| 0.403249 | 0.166275 | | | |
| 0.549897 | 0.013012 | | *Variable 1* | *Variable 2* |
| 0.515156 | 0.19465 | Mean | 0.49092382 | 0.61498223 |
| 0.160181 | 0.101519 | Variance | 0.1810696 | 0.54459685 |
| 0.198179 | 0.837068 | Observations | 11 | 13 |
| 0.687517 | 0.116898 | Pooled Varian | 0.37935719 | |
| 0.058422 | 0.305804 | Hypothesized | 0 | |
| 0.454039 | 2.08066 | df | 22 | |
| 1.60372 | 1.05122 | t Stat | -0.4916595 | |
| 0.625722 | 2.02541 | P(T<=t) one-ta | 0.31391488 | |
| 0.14408 | 1.0136 | t Critical one- | 1.71714437 | |
| | 0.036652 | P(T<=t) two-ta | 0.62782977 | |
| | 0.052001 | t Critical two-t | 2.07387307 | |
| **CD16** | | | | |

| R | NR | t-Test: Two-Sample Assuming Equal Variar | | |
|---|---|---|---|---|
| 0.450149 | 0.52933 | | | |
| 1.50816 | 0.320935 | | *Variable 1* | *Variable 2* |
| 2.87714 | 0.206143 | Mean | 1.58730273 | 0.79601438 |
| 0.247551 | 0.134237 | Variance | 3.12720935 | 1.52400645 |
| 0.525514 | 0.39955 | Observations | 11 | 13 |
| 1.61127 | 0.149053 | Pooled Varian | 2.25273504 | |
| 0.098446 | 0.188254 | Hypothesized | 0 | |
| 1.09866 | 4.11094 | df | 22 | |
| 6.35834 | 0.024784 | t Stat | 1.28689202 | |
| 1.49756 | 1.2308 | P(T<=t) one-ta | 0.1057542 | |
| 1.18754 | 2.71898 | t Critical one- | 1.71714437 | |
| | 0.172412 | P(T<=t) two-ta | 0.2115084 | |
| | 0.162769 | t Critical two- | 2.07387307 | |
| **CK** | | | | |

| R | NR | t-Test: Two-Sample Assuming Equal Variar | | |
|---|---|---|---|---|
| 3.70364 | 5.66575 | | | |
| 6.33536 | 9.11698 | | *Variable 1* | *Variable 2* |
| 4.77248 | 5.31388 | Mean | 5.90309 | 4.50508615 |
| 1.20097 | 3.28152 | Variance | 14.9347595 | 4.80849013 |
| 6.6509 | 2.82256 | Observations | 11 | 13 |
| 3.23678 | 1.91094 | Pooled Varian | 9.41133982 | |
| 11.6325 | 3.9655 | Hypothesized | 0 | |
| 14.3784 | 7.77654 | df | 22 | |
| 5.2587 | 4.03241 | t Stat | 1.11235916 | |
| 3.76877 | 6.30539 | P(T<=t) one-ta | 0.1389958 | |
| 3.99549 | 2.65717 | t Critical one- | 1.71714437 | |
| | 3.05829 | P(T<=t) two-ta | 0.2779916 | |
| | 2.65919 | t Critical two- | 2.07387307 | |
| **SMA** | | | | |

| R | NR | t-Test: Two-Sample Assuming Equal Variar | | |
|---|---|---|---|---|
| 0.743305 | 0.748031 | | | |
| 0.998669 | 2.21944 | | *Variable 1* | *Variable 2* |
| 1.6394 | 0.496713 | Mean | 1.08155555 | 1.07305454 |
| 0.404291 | 1.05138 | Variance | 0.34158029 | 0.93302335 |
| 2.28837 | 1.12569 | Observations | 11 | 13 |
| 1.00109 | 0.501058 | Pooled Varian | 0.6641856 | |
| 1.21887 | 0.221269 | Hypothesized | 0 | |
| 1.57718 | 1.8876 | df | 22 | |
| 0.866895 | 0.45847 | t Stat | 0.02546174 | |
| 0.228017 | 3.58635 | P(T<=t) one-ta | 0.48995811 | |

| | | | |
|---|---|---|---|
| 0.931024 | 0.8666 | t Critical one- | 1.71714437 |
| | 0.572989 | P(T<=t) two-ta | 0.97991621 |
| | 0.214119 | t Critical two-t | 2.07387307 |
