## Supplementary Table 6 for "Immune Cell Densities Predict Response to Immune Checkpoint-Blockade in Head and Neck Cancer"

Supplementary Table 6. Percentage of contribution for each marker to Tertiary Lymphoid Structures

| Group | Sample | CD16 | SMA | Ki67 | CD56 | CK | CD19 | CD163 | PD-1 | LAMP3 | CD8 | CD20 | CD4 | # of TLS | Average TLS area (µm^2) | Smallest (µm^2) | Largest (µm^2) | Average TLS Perimeter (µm) |
|---|---|---|---|---|---|---|---|---|---|---|---|---|---|---|---|---|---|---|
| NR | ICI-14_G7 | 50867 | 9794 | 9829 | 12169 | 7457 | 207 | 54138 | 57717 | 371 | 21564 | 61012 | 101334 | 22 | 688686 | 9654.57 | 3035736.2 | 25215.454 |
| | % | 13.162 | 2.534 | 2.543 | 3.149 | 1.93 | 0.054 | 14.009 | 14.935 | 0.096 | 5.57989 | 15.79 | 26.221 | | | | | |
| | ICI-9_P8 | 1 | 14 | 80 | 65 | 10 | 559 | 84 | 389 | 22 | 289 | 990 | 1653 | 2 | 147286 | 52797.3 | 241774.94 | 6871.9999 |
| | % | 0.0241 | 0.337 | 1.925 | 1.564 | 0.24 | 13.45 | 2.0212 | 9.36 | 0.5294 | 6.9538 | 23.82 | 39.774 | | | | | |
| | ICI-23 | 32 | 113 | 152 | 115 | 106 | 560 | 58 | 187 | 11 | 65 | 1206 | 702 | 1 | 145982 | 145982 | 145981.7 | 3312 |
| | % | 0.9676 | 3.417 | 4.596 | 3.477 | 3.21 | 16.93 | 1.7539 | 5.6547 | 0.3326 | 1.96553 | 36.47 | 21.228 | | | | | |
| | ICI-25_140 | 164 | 7 | 76 | 0 | 0 | 0 | 6 | 14 | 5 | 176 | 117 | 298 | 1 | 66572 | 66572 | 66571.96 | |
| | % | 19.003 | 0.811 | 8.806 | 0 | 0 | 0 | 0.6952 | 1.6222 | 0.5794 | 20.394 | 13.56 | 34.531 | | | | | |
| | ICI-26 | NONE | | | | | | | | | | | | 0 | NONE | | | |
| | ICI-28 | NONE | | | | | | | | | | | | 0 | NONE | | | |
| | ICI-30 | 355 | 142 | 230 | 243 | 11 | 16 | 1012 | 1047 | 263 | 864 | 801 | 1583 | 5 | 67831 | 8845.59 | 145016.4 | 3184 |
| | % | 5.4058 | 2.162 | 3.502 | 3.7 | 0.17 | 0.244 | 15.41 | 15.943 | 4.0049 | 13.1567 | 12.2 | 24.105 | | | | | |
| | ICI-37 | 714 | 1082 | 1214 | 1303 | 199 | 2 | 2281 | 2798 | 281 | 5322 | 5055 | 12185 | 16 | 118076 | 36709.2 | 482411.1 | 5357.2499 |
| | % | 2.2013 | 3.336 | 3.743 | 4.017 | 0.61 | 0.006 | 7.0323 | 8.6262 | 0.8663 | 16.4077 | 15.58 | 37.566 | | | | | |
| | CTC-21_20 | NONE | | | | | | | | | | | | 0 | NONE | | | |
| | S-69_M3 | 18217 | 11003 | 4439 | 5489 | 1410 | 1379 | 25302 | 19857 | 129 | 3957 | 11300 | 40896 | 10 | 453668 | 137530 | 859688.3 | 26316.4 |
| | % | 12.706 | 7.674 | 3.096 | 3.828 | 0.98 | 0.962 | 17.647 | 13.849 | 0.09 | 2.75984 | 7.881 | 28.523 | | | | | |
| | ICI-33 | 2332 | 998 | 414 | 208 | 4843 | 99 | 1156 | 590 | 277 | 4676 | 2693 | 6370 | 13 | 50967 | 15688.1 | 175578.3 | 2548 |
| | % | 9.4581 | 4.048 | 1.679 | 0.844 | 19.6 | 0.402 | 4.6885 | 2.3929 | 1.1235 | 18.965 | 10.92 | 25.835 | | | | | |
| | ICI-5 | 17 | 1488 | 310 | 13 | 128 | 2 | 0 | 16 | 11 | 1711 | 2458 | 2719 | 1 | 346103 | 346103 | 346103.2 | 8919.999 |
| | % | 0.1916 | 16.77 | 3.494 | 0.147 | 1.44 | 0.023 | 0 | 0.1803 | 0.124 | 19.2832 | 27.7 | 30.644 | | | | | |
| | ICI-13_B1_ | NONE | | | | | | | | | | | | 0 | NONE | | | |
| R | ICI-3 | NONE | | | | | | | | | | | | 0 | NONE | | | |
| | ICI-9_1405 | NONE | | | | | | | | | | | | 0 | NONE | | | |
| | ICI-11 | 1932 | 7776 | 3329 | 3710 | 6408 | 23954 | 1867 | 13321 | 753 | 8215 | 49576 | 28730 | 24 | 254977 | 22211.7 | 690703.6 | 9311.6667 |
| | % | 1.2917 | 5.199 | 2.226 | 2.48 | 4.28 | 16.02 | 1.2482 | 8.9061 | 0.5034 | 5.49237 | 33.15 | 19.208 | | | | | |
| | ICI-16 | 1268 | 2709 | 2327 | 1548 | 2120 | 10685 | 3858 | 17547 | 4004 | 9805 | 20183 | 26777 | 9 | 468404 | 62174.8 | 1782309 | 14162.222 |
| | % | 1.2331 | 2.634 | 2.263 | 1.505 | 2.06 | 10.39 | 3.7518 | 17.064 | 3.8938 | 9.53506 | 19.63 | 26.04 | | | | | |
| | ICI-18_X-0 | 171304 | 18865 | 26883 | 15947 | 6469 | 49458 | 111436 | 104300 | 970 | 115114 | 1E+05 | 151745 | 25 | 1E+06 | 25131.6 | 5986057 | 41223.52 |
| | % | 19.301 | 2.126 | 3.029 | 1.797 | 0.73 | 5.573 | 12.556 | 11.752 | 0.1093 | 12.9702 | 12.96 | 17.098 | | | | | |
| | ICI-19_A5 | NONE | | | | | | | | | | | | 0 | | | | |
| | CTC-13 | 14212 | 5518 | 4322 | 1258 | 608 | 3508 | 3031 | 5492 | 476 | 3935 | 15994 | 21720 | 19 | 186153 | 34631.2 | 869879.44 | 7022.5262 |
| | % | 17.749 | 6.891 | 5.398 | 1.571 | 0.76 | 4.381 | 3.7852 | 6.8587 | 0.5945 | 4.9142 | 19.97 | 27.125 | | | | | |
| | CTC-15 | 4703 | 1708 | 6555 | 1234 | 82 | 9942 | 3547 | 3337 | 510 | 4290 | 10404 | 18171 | 6 | 419300 | 117804 | 1728599.2 | 14167.333 |
| | % | 7.2934 | 2.649 | 10.17 | 1.914 | 0.13 | 15.42 | 5.5007 | 5.175 | 0.7909 | 6.65292 | 16.13 | 28.18 | | | | | |
| | CTC-18 | 32805 | 3846 | 12089 | 1218 | 253 | 2362 | 14468 | 7341 | 1154 | 24207 | 38356 | 21997 | 18 | 387188 | 12450.3 | 2873445.2 | 16566.889 |
| | % | 20.491 | 2.402 | 7.551 | 0.761 | 0.16 | 1.475 | 9.0371 | 4.5854 | 0.7208 | 15.1203 | 23.96 | 13.74 | | | | | |
| | CTC-19_14 | 10264 | 3607 | 4988 | 326 | 325 | 869 | 1921 | 1336 | 253 | 14172 | 19204 | 16617 | 10 | 410249 | 36151 | 1179536 | 19989.6 |
| | % | 13.892 | 4.882 | 6.751 | 0.441 | 0.44 | 1.176 | 2.6001 | 1.8083 | 0.3424 | 19.1819 | 25.99 | 22.491 | | | | | |
| | CTC-23 | NONE | | | | | | | | | | | | 0 | NONE | | | |

| | CD16 (Imne \| Opal 520 Positive | SMA (Imne \| Opal 540 | Ki67 (Imne \| Opal 570 | CD56 (Imne \| Opal 620 | CK (Imne \| Opal 650 | CD19 (Imne \| Opal 690 | CD163 (TLS \| Opal 690 Positive | PD-1 (TLS \| Opal 520 Positive | LAMP3 (TLS \| Opal 540 Positive | CD8 (TLS \| Opal 570 Positive Cells) | CD20 (TLS \| Opal 620 | CD4 (TLS \| Opal 650 Positive |
|---|---|---|---|---|---|---|---|---|---|---|---|---|
| **Total%** | 14.522 | 3.225 | 3.628 | 2.106 | 1.43 | 4.866 | 10.528 | 11.051 | 0.4457 | 10.2558 | 16.64 | 21.299 |
