## Supplementary Table 5 for "Immune Cell Densities Predict Response to Immune Checkpoint-Blockade in Head and Neck Cancer"

Supplementary Table 5. Tertiary Lymphoid structures, mIF and pathologist

| ID | Response | Tissue size (mm2) | Pathologist | mIF (greater or equal to 5 LAMP3 and 50 CD20) mature | miF/size | Average distance of TLS to tumor area |
|---|---|---|---|---|---|---|
| ICI-19_A4 | R | 154.3057 | 0 | 0 | 0 | NA |
| ICI-28 | NR | 89.03552 | 0 | 0 | 0 | NA |
| CTC-13 | R | 54.02518 | 3 | 19 | 0.35168786 | 171.88 |
| CTC-15 | R | 31.4382 | 2 | 6 | 0.19085062 | 130.69 |
| **CTC-18*** | **R** | **243.29583** | 4 | 18 | 0.07398401 | 98.97 |
| CTC-23 | R | 33.83882 | 0 | 0 | 0 | NA |
| ICI-3 | R | 57.86318 | 0 | 0 | 0 | NA |
| ICI-5 | NR | 20.13121 | 0 | 1 | 0.04967411 | 8.43 |
| ICI-9_P8** | NR | 63.81032 | 0 | 2 | 0.03134289 | 159.05 |
| ICI-11 | R | 150.87018 | 15 | 24 | 0.15907716 | 57.35 |
| ICI-16*** | R | 127.9709 | 1 | 9 | 0.07032849 | 57.14 |
| ICI-23**** | NR | 83.82644 | 0 | 1 | 0.01192941 | 157.3 |
| ICI-26 | NR | 93.46102 | 0 | 0 | 0 | NA |
| ICI-30 | NR | 127.63196 | 4 | 5 | 0.03917514 | 122.68 |
| ICI-33 | NR | 155.79375 | 0 | 13 | 0.08344366 | 125.1 |
| ICI-37 | NR | 170.15154 | 1 | 16 | 0.09403382 | 115.68 |
| S-69_M3 | NR | 84.62711 | 4 | 10 | 0.11816544 | 108.83 |
| 1340 ICI-18_X-07_A6 | R | 119.48057 | 10 | 25 | 0.20923904 | 114.89 |
| 1430 CTC-19* | R | 127.32778 | 5 | 10 | 0.07853746 | 208.72 |
| **1405 ICI-9*** ** | **R** | **158.81167** | 0 | 0 | 0 | NA |
| **1431 CTC-21**** | **NR** | **164.11794** | 0 | 0 | 0 | NA |
| 1309 ICI-13_B1 | NR | 55.05734 | 0 | 0 | 0 | NA |
| **1408 ICI-25******** | **NR** | **119.19083** | 1 | 1 | 0.00838991 | 187.79 |
| ICI-14_G7 | NR | 205.21322 | 12 | 22 | 0.10720557 | 120.3 |

CTC-18 and CTC-19 are replicates from the same patient

ICI-9_P8 and CTC-21 are replicates from the same patient

ICI-16 and ICI-9_1405 are replicates from the same patient

ICI-23 and ICI-25 are replicates from the same patient

Excluded from OS and PFS analysis: 1430_CTC-19, ICI-9_P8, ICI-16, ICI-23 (The replicate with largest tissue analyzed was used instead).
